# Violence Exposure and Mental Health Problems Among School-Aged Children in a South African Birth Cohort

**DOI:** 10.64898/2026.04.20.26351289

**Authors:** Megan Bailey, Gemma Hammerton, Graeme Fairchild, Lucinda Tsunga, Nadia Hoffman, Tiffany Burd, Richard Shadwell, Andrea Danese, Cherie Armour, Heather J Zar, Dan J Stein, Kirsten Donald, Sarah L Halligan

## Abstract

**Objective:** There is little longitudinal research investigating links between violence exposure and mental disorders among children in low- and middle-income countries (LMICs), despite high rates of violence. We examined cross-sectional and longitudinal violence-mental health associations among children in a large South African birth cohort, the Drakenstein Child Health Study, including direct clinical interviews capturing children’s mental disorders.

**Method:** In this birth cohort (*N*=974), we assessed lifetime violence exposure and four subtypes (witnessed community, community victimization, witnessed domestic, domestic victimization) at ages 4.5 and 8-years via caregiver reports. At 8-years, caregivers completed the Child Behaviour Checklist; and psychiatric disorders were assessed using the Mini-International Neuropsychiatric Interview for Children and Adolescents, a self-report measure. We tested for associations using linear/logistic regressions, adjusted for confounders.

**Results:** Most children (91%) had experienced violence by 8-years. Cross-sectionally, total violence exposure was associated with total (B =0.49 [95% CI 0.32, 0.66]), internalizing (0.32 [0.17, 0.47]), and externalizing problems (0.46 [0.31, 0.61]), and with increased odds of disorder at 8 years (aOR=1.09 [1.05, 1.13]). Longitudinally, total violence exposure up to 4.5-years was associated with total (B=0.27 [0.03, 0.52]), internalizing (0.24 [0.04. 0.44]), and externalizing scores (0.23 [0.008, 0.45]) at 8-years, but not with increased risk of psychiatric disorders. The strongest and most consistent associations were observed for domestic versus community violence subtypes.

**Conclusion:** Our strong cross-sectional but weaker longitudinal findings suggest that recent violence exposures may be more critical than early exposures for children’s mental health. Longitudinal exploration of other violence-affected LMIC populations is urgently needed.

## INTRODUCTION

Globally, it is estimated that around half of all children and adolescents are exposed to some form of violence each year,^1^ including maltreatment (e.g., physical and sexual abuse), caregiver intimate partner violence, and community violence. Systematic reviews and meta-analyses have consistently linked these forms of violence exposure to adverse mental health outcomes throughout the lifespan,^2–4^ but especially among children and adolescents.^5–7^ Additionally, exposure to multiple forms of violence, or polyvictimisation, is linked to an increased risk of mental health problems, with evidence of a dose-response relationship such that exposure to more types of violence is associated with greater risk of mental health problems.^8,9^ However, this evidence base still has several key limitations.

First, studies have primarily used cross-sectional designs, in which violence exposure and mental health outcomes are reported at the same time, severely limiting the ability to draw causal conclusions and potentially being biased by inaccurate memory recall.^10^ Second, research has primarily been conducted in high-income countries, providing limited insight into the experiences of the vast majority of children and adolescents worldwide who live in low- and middle-income countries (LMICs).^11^ This omission is particularly problematic given that LMIC populations typically face greater risk of violence exposure and poorer access to healthcare services than those in high-income countries.^12,13^ Notably, in a recent scoping review, only 25.9% (15 of 58) of studies examining violence-mental health associations among LMIC children had longitudinal designs.^14^ The vast majority of studies focused on a single violence type (mostly maltreatment) and used a single informant to assess violence exposure and mental health symptoms. No longitudinal study examined clinical diagnoses as the outcome, with just one cross-sectional study assessing clinical diagnoses via caregiver interviews. Given these limitations, longitudinal research is critical to advance our understanding of violence exposure as an important influence on the mental health of the 90% of children globally who reside in LMICs, a population that is expected to continue rising over the next several decades, especially in Africa.^15^

We addressed these limitations by investigating associations between childhood violence exposure and mental health problems in an ongoing South African birth cohort, the Drakenstein Child Health Study (DCHS). Recent research found that >75% of this cohort was exposed to violence by age 6 years,^16^ and established longitudinal associations between caregiver-reported violence and internalizing/externalizing symptoms by age 5.^17^ We extended these findings to examine mental health in children aged 8 years, an age at which school-based victimization becomes an important potential contributor to violence in this population,^18,19^ and the prevalence of psychological disorders begins to increase.^20,21^ Crucially, we used diagnostic data obtained from structured clinical interviews with children at age 8, alongside caregiver reports of children’s symptoms and violence exposure, obtaining child-reported disorder prevalence figures for the first time and limiting potential for single informant bias.^22^ We examined cross-sectional violence-mental health associations at 8 years, and additionally tested for persistent effects of early violence exposure (up to 4.5 years) on 8-year mental health outcomes. We also investigated whether there were sex differences in these associations, given that previous research reported differential effects of violence subtypes in boys and girls in South Africa.^23^

## METHODS

### Study design and population

Between 2012 and 2015, the DCHS recruited pregnant women at 20-28 weeks’ gestation from two public sector primary health care clinics in the Drakenstein sub-district of the Cape Winelands, Western Cape, South Africa.^24^ Eligible pregnant women were at least 18 years old. Mother-child dyads attended follow-up visits from birth throughout childhood. The study was approved by the Faculty of Health Sciences, Human Research Ethics Committee, University of Cape Town (401/2009), Stellenbosch University (N12/02/0002), and the Western Cape Provincial Health Research committee (2011RP45), with all standard ethical procedures relating to informed consent, assent, and risk management adhered to throughout.

### Measures

#### Childhood violence exposure

Child lifetime violence exposure was assessed at ages 4.5 and 8 years using the caregiver-report Child Exposure to Community Violence Checklist (CECV).^25^ The CECV includes 35 items, answered on a 4-point Likert scale (ranging from 0 “never” to 3 “many times”). An adapted version of the CECV, which has shown good reliability in previous studies,^23,26^ was used to better fit the South African context. Responses were summed to generate a total violence exposure score (range=0-105). We also examined violence exposure across four subscales that were used in previous research with this cohort^16^: witnessed community violence (10 items), community victimization (8 items), witnessed domestic violence (6 items), and domestic victimization (11 items). Finally, we captured polyvictimisation by coding the number of subscales in which a child had experienced an event, with a maximum score of 4 indicating that the child was exposed to events across all four types of violence. Likelihood-ratio tests suggested linearity in polyvictimisation-psychopathology associations and the suitability of treating polyvictimisation as a numeric, rather than a categorical, exposure (see Appendix 1).

#### Child psychopathology

Child mental health was assessed at age 8 years using the caregiver-report Child Behaviour Checklist (CBCL).^27^ The CBCL assesses child emotional and behavioral problems in the past 6-months and is comprised of 113 items, answered on a 3-point Likert scale (0 “not true”, 1 “somewhat or sometimes true”, and 2 “very true or often true”). We utilized total problems (sum of all 113 items; range=0-226) and two second-order categories of syndromes: internalizing problems (sum of 36 items; range=0-72) and externalizing problems (sum of 24 items; range=0-48). Consistent with standard practice, raw scores were converted into *t*-scores with a mean of 50 and a standard deviation of 10 (based on norm-referenced scores from the US).

Additionally, current child psychiatric disorders were assessed at 8 years via structured clinical interviews with the child using the Mini-International Neuropsychiatric Interview for Children and Adolescents (MINI-KID).^28^ Ten diagnoses were assessed: generalized anxiety disorder, obsessive compulsive disorder, posttraumatic stress disorder, separation anxiety disorder, social anxiety disorder, specific phobia, major depressive disorder, attention-deficit/hyperactivity disorder (ADHD), conduct disorder, and oppositional defiant disorder. We used an ‘any disorder’ binary variable which captured whether the child met criteria for at least one diagnosis.

#### Confounders

Child sex and ancestry were captured at birth. Household income and maternal educational background, employment status, and marital status were assessed at enrolment. Maternal depression was assessed at enrolment using the self-report Edinburgh Postnatal Depression Scale (EPDS).^29^ Maternal alcohol use and smoking during pregnancy were assessed at enrolment using the Alcohol, Smoking and Substance Involvement Screening Test (ASSIST),^30^ and during postpartum assessments at 3-6 weeks and 2 years. We used binary composite indicators to represent any reported prenatal alcohol use and smoking. Maternal HIV status during pregnancy was confirmed by routine testing and further re-testing of HIV- negative mothers every 12 weeks. HIV status reviews of mothers and children were conducted at birth, age 6 weeks, and every 6-months thereafter until age 18 months or at cessation of breastfeeding if this occurred after 18 months (for further details see Wedderburn et al., 2019).^31^ Two children who tested positive for HIV were excluded from the analyses.

We used a dichotomous variable to capture HIV exposed versus unexposed uninfected children. Child age at the 8-year CECV assessment was also included as a confounder in our cross-sectional models given variation in child age at this follow-up due to the COVID pandemic (mean=8.46 years [SD 0.50], range=7.77-10.58 years). Finally, pre-existing child psychopathology was assessed at 3.5 years using the CBCL.

### Procedure

The CECV, CBCL, and MINI-KID were translated from English to Afrikaans and isiXhosa by teams consisting of at least 3 mother-tongue speakers for each language. Translations were cross-checked by community-based study staff who were fluent in the relevant languages to ensure that a suitable dialect was used, and consensus meetings were also held. Data for all three measures were collected and managed using REDCap electronic data capture tools.^32,33^ Trained research assistants with a background in a relevant field (social work, psychology) administered the CECV and MINI-KID at an in-person clinic visit. The CBCL was administered in person or via telephone (depending on caregiver availability) by trained fieldworkers. Typically, in the DCHS, the CBCL and the CECV are administered at different clinic visits within a period of ∼6 months. Due to the COVID-19 pandemic extending the 8-year follow-up period, the CBCL was re-administered at the same clinic visit as the CECV if the CBCL had previously been conducted more than 6-months prior to the CECV assessment. Finally, a caregiver was eligible to complete the CECV and CBCL if they lived with or cared for the child for ≥3 days per week.

### Data analysis

We pre-registered our analyses on the Open Science Framework (https://doi.org/10.17605/OSF.IO/GZE2U; see Appendix 2). Analyses were conducted using Stata Version 18. Multiple imputation by chained equations with 50 imputed datasets was used to address missing data (see Appendix 3); the findings presented here are based on imputed data (see Appendix 4 for complete case analyses).

We used linear regression analyses to examine associations between violence exposure scores (total, subscales, and polyvictimisation) and CBCL scores (total, internalizing, and externalizing). Examination of model diagnostics revealed some evidence of heteroscedasticity; we therefore used robust standard errors in all analyses (see Appendix 5 for further discussion of model diagnostics). We also used Firth’s penalized likelihood regression to examine associations between violence exposure scores and any psychiatric disorder (due to a low prevalence of psychiatric disorders in the sample). Cross-sectional analyses examined violence exposure scores at 8 years in relation to concurrent psychopathology. Longitudinal analyses examined violence exposure scores at 4.5 years in relation to psychopathology at 8 years. For each analysis, we conducted three models: unadjusted, adjusted for baseline confounders (and child age for cross-sectional models), and finally a model that additionally adjusted for child psychopathology at age 3.5 (to explore whether childhood violence exposure predicted psychopathology over and above any early childhood psychopathology). Finally, in sensitivity analyses, we examined whether violence-psychopathology associations differed by sex.

## RESULTS

At birth, 1137 mothers were enrolled in the study, with 1143 live births. Of these 1143 children, 977 (85.5%) are currently active in the cohort, and 974 were included in our analysis sample (85.2% of the total sample; *n*=2 children were excluded due to testing HIV positive and *n*=1 child was excluded due to incomplete ancestry data). Compared to our analysis sample, mothers of inactive and excluded children were less likely to have smoked during pregnancy (odds ratio (OR)=0.65 [95% CI 0.44, 0.97], *p*=0.034), were more likely to have completed secondary or any tertiary education (compared to primary or some secondary education; 1.70 [1.22, 2.36], *p*=0.002), and had greater odds of a higher family income (>R5000/m compared to <R1000/m; 2.51 [1.55, 4.04], *p*<0.001; Appendix 6).

Sample characteristics are presented in Table 1. As previously reported,^16^ at 4.5 years, 75.4% of children had been exposed to any type of violence, with witnessing community violence being the most prevalent type (65.6%). At 8 years, 91.1% of children had been exposed to any type of violence. Witnessing community violence remained the most prevalent type of violence exposure (74.7%) and witnessing domestic violence the least prevalent (24.3%). Just over half of children experienced more than one violence type by age 8 (53.6%; for the prevalence of individual events see Appendix 7). For child psychopathology at age 8, mean *t*-scores on the CBCL were less than 50, indicating lower-than-average scores compared to the American reference norms, with 11.4% of children meeting criteria for at least one psychiatric disorder. Externalizing disorders were more prevalent (8.0%) than internalizing disorders (3.7%), and ADHD (6.8%), conduct disorder (3.0%), oppositional defiant disorder (2.3%), and specific phobias (2.0%) were the most common diagnoses (for the prevalence of all other disorders see Appendix 8).

**Table 1.**
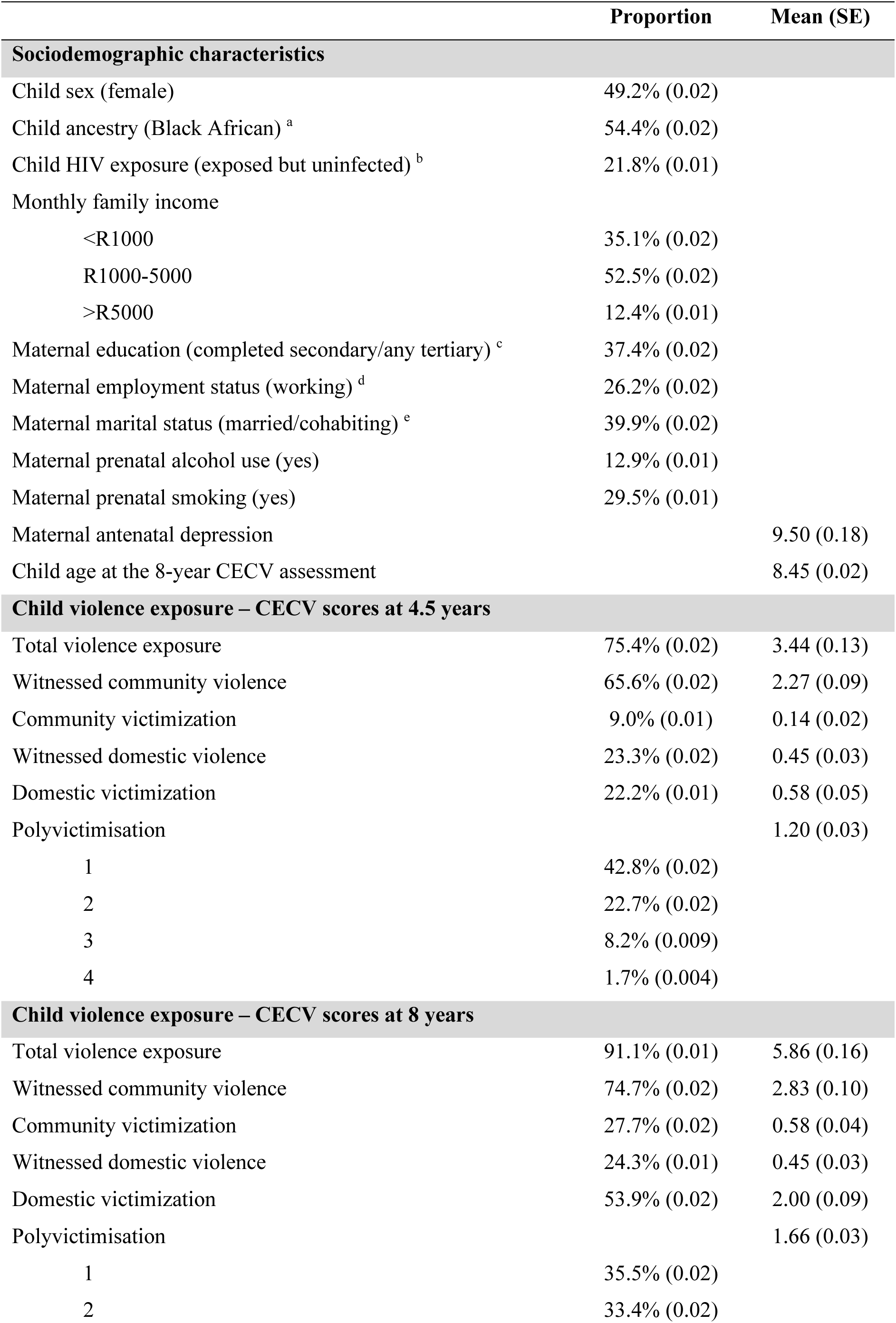
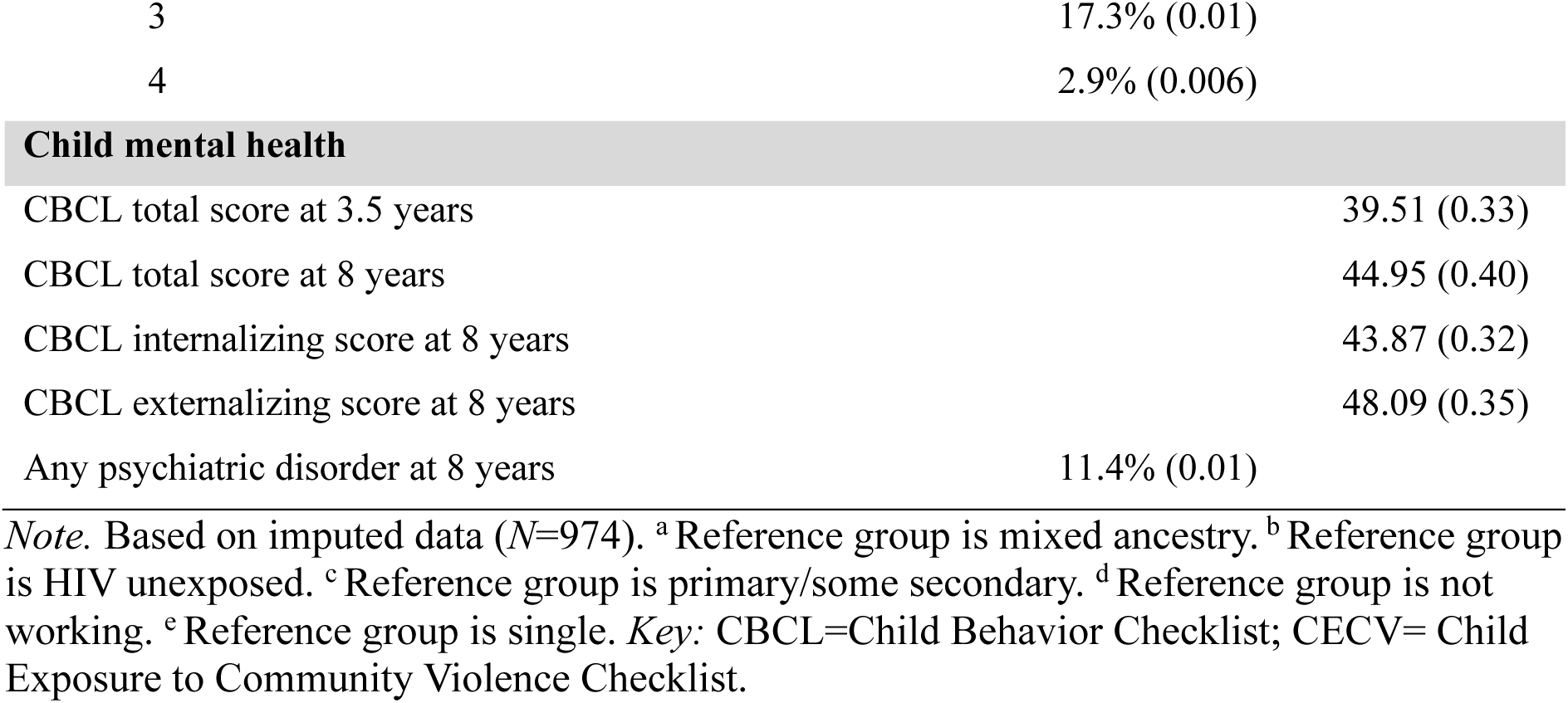
Sociodemographic characteristics, violence exposure, and mental health descriptives.

### Cross-sectional associations between violence exposure at 8 years and child psychopathology at 8 years

Table 2 presents the unadjusted and adjusted associations between childhood violence exposure scores at 8 years and mental health problem scores at 8 years. In fully adjusted analyses, total violence exposure scores at 8 years were positively associated with total problem (B for each one-unit increase in violence score=0.49 [95% CI 0.32, 0.66], *p*<0.001), internalizing (0.32 [0.17, 0.47], *p*<0.001), and externalizing scores (0.46 [0.31, 0.61], *p*<0.001). Scores on all three CBCL scales were also positively associated with subscale scores for witnessing community violence, witnessing domestic violence, and domestic victimization, but not community victimization. Polyvictimisation was positively associated with higher total problem and externalizing scores only (see Table 2).

**Table 2.**
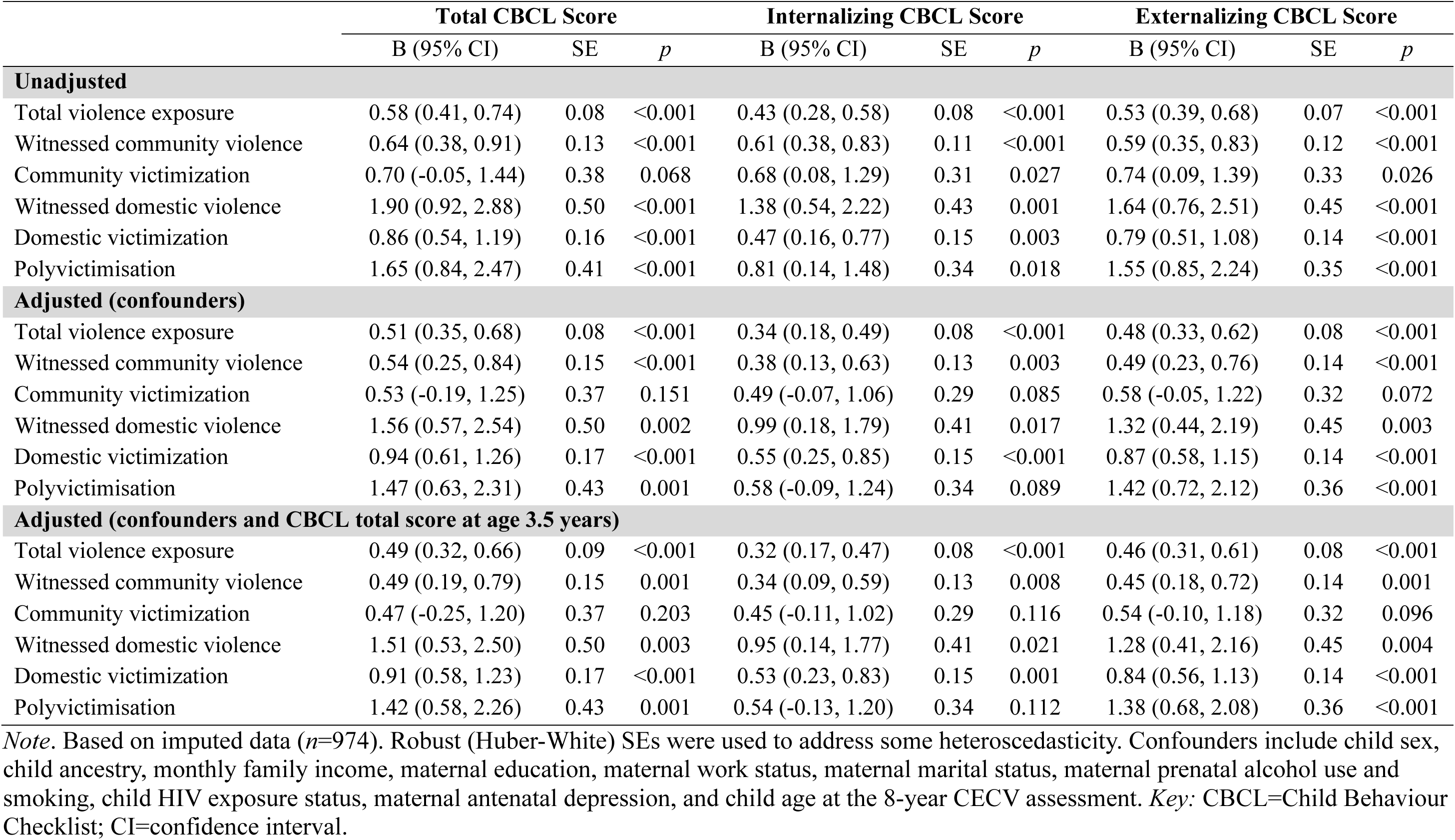
Cross-sectional associations between violence exposure score at age 8 years and mental health problems at age 8 years.

|  | Total CBCL Score |  |  | Internalizing CBCL Score |  |  | Externalizing CBCL Score |  |  |
| --- | --- | --- | --- | --- | --- | --- | --- | --- | --- |
|  | B (95% CI) | SE | *p* | B (95% CI) | SE | *p* | B (95% CI) | SE | *p* |
| **Unadjusted** |  |  |  |  |  |  |  |  |  |
| Total violence exposure | 0.58 (0.41, 0.74) | 0.08 | <0.001 | 0.43 (0.28, 0.58) | 0.08 | <0.001 | 0.53 (0.39, 0.68) | 0.07 | <0.001 |
| Witnessed community violence | 0.64 (0.38, 0.91) | 0.13 | <0.001 | 0.61 (0.38, 0.83) | 0.11 | <0.001 | 0.59 (0.35, 0.83) | 0.12 | <0.001 |
| Community victimization | 0.70 (-0.05, 1.44) | 0.38 | 0.068 | 0.68 (0.08, 1.29) | 0.31 | 0.027 | 0.74 (0.09, 1.39) | 0.33 | 0.026 |
| Witnessed domestic violence | 1.90 (0.92, 2.88) | 0.50 | <0.001 | 1.38 (0.54, 2.22) | 0.43 | 0.001 | 1.64 (0.76, 2.51) | 0.45 | <0.001 |
| Domestic victimization | 0.86 (0.54, 1.19) | 0.16 | <0.001 | 0.47 (0.16, 0.77) | 0.15 | 0.003 | 0.79 (0.51, 1.08) | 0.14 | <0.001 |
| Polyvictimisation | 1.65 (0.84, 2.47) | 0.41 | <0.001 | 0.81 (0.14, 1.48) | 0.34 | 0.018 | 1.55 (0.85, 2.24) | 0.35 | <0.001 |
| **Adjusted (confounders)** |  |  |  |  |  |  |  |  |  |
| Total violence exposure | 0.51 (0.35, 0.68) | 0.08 | <0.001 | 0.34 (0.18, 0.49) | 0.08 | <0.001 | 0.48 (0.33, 0.62) | 0.08 | <0.001 |
| Witnessed community violence | 0.54 (0.25, 0.84) | 0.15 | <0.001 | 0.38 (0.13, 0.63) | 0.13 | 0.003 | 0.49 (0.23, 0.76) | 0.14 | <0.001 |
| Community victimization | 0.53 (-0.19, 1.25) | 0.37 | 0.151 | 0.49 (-0.07, 1.06) | 0.29 | 0.085 | 0.58 (-0.05, 1.22) | 0.32 | 0.072 |
| Witnessed domestic violence | 1.56 (0.57, 2.54) | 0.50 | 0.002 | 0.99 (0.18, 1.79) | 0.41 | 0.017 | 1.32 (0.44, 2.19) | 0.45 | 0.003 |
| Domestic victimization | 0.94 (0.61, 1.26) | 0.17 | <0.001 | 0.55 (0.25, 0.85) | 0.15 | <0.001 | 0.87 (0.58, 1.15) | 0.14 | <0.001 |
| Polyvictimisation | 1.47 (0.63, 2.31) | 0.43 | 0.001 | 0.58 (-0.09, 1.24) | 0.34 | 0.089 | 1.42 (0.72, 2.12) | 0.36 | <0.001 |
| **Adjusted (confounders and CBCL total score at age 3.5 years)** |  |  |  |  |  |  |  |  |  |
| Total violence exposure | 0.49 (0.32, 0.66) | 0.09 | <0.001 | 0.32 (0.17, 0.47) | 0.08 | <0.001 | 0.46 (0.31, 0.61) | 0.08 | <0.001 |
| Witnessed community violence | 0.49 (0.19, 0.79) | 0.15 | 0.001 | 0.34 (0.09, 0.59) | 0.13 | 0.008 | 0.45 (0.18, 0.72) | 0.14 | 0.001 |
| Community victimization | 0.47 (-0.25, 1.20) | 0.37 | 0.203 | 0.45 (-0.11, 1.02) | 0.29 | 0.116 | 0.54 (-0.10, 1.18) | 0.32 | 0.096 |
| Witnessed domestic violence | 1.51 (0.53, 2.50) | 0.50 | 0.003 | 0.95 (0.14, 1.77) | 0.41 | 0.021 | 1.28 (0.41, 2.16) | 0.45 | 0.004 |
| Domestic victimization | 0.91 (0.58, 1.23) | 0.17 | <0.001 | 0.53 (0.23, 0.83) | 0.15 | 0.001 | 0.84 (0.56, 1.13) | 0.14 | <0.001 |
| Polyvictimisation | 1.42 (0.58, 2.26) | 0.43 | 0.001 | 0.54 (-0.13, 1.20) | 0.34 | 0.112 | 1.38 (0.68, 2.08) | 0.36 | <0.001 |

*Note.* Based on imputed data ( $n=974$ ). Robust (Huber-White) SEs were used to address some heteroscedasticity. Confounders include child sex, child ancestry, monthly family income, maternal education, maternal work status, maternal marital status, maternal prenatal alcohol use and smoking, child HIV exposure status, maternal antenatal depression, and child age at the 8-year CECV assessment. *Key:* CBCL=Child Behaviour Checklist; CI=confidence interval.

Table 3 presents the unadjusted and adjusted associations between childhood violence exposure scores at age 8 years and the presence of any psychiatric disorder at age 8 years. In fully adjusted analyses, each one-unit increase in the total violence exposure score was associated with a 9% increase in the odds of any psychiatric disorder at age 8 years (adjusted OR=1.09 [95% CI 1.05, 1.13], *p*<0.001). Community victimization, witnessing domestic violence, domestic victimization, and polyvictimisation were also each associated with increased odds of disorder at age 8 years, but the effect of witnessing community violence was weaker (1.07 [0.99, 1.15], *p*=0.080; see Table 3).

**Table 3.** Cross-sectional associations between violence exposure score at age 8 years and psychiatric disorders at age 8 years.

|  | Any disorder |  |
| --- | --- | --- |
|  | OR (95% CI) | *p* |
| **Unadjusted** |  |  |
| Total violence exposure | 1.11 (1.07, 1.15) | <0.001 |
| Witnessed community violence | 1.08 (1.01, 1.15) | 0.035 |
| Community victimization | 1.29 (1.13, 1.48) | <0.001 |
| Witnessed domestic violence | 1.57 (1.29, 1.90) | <0.001 |
| Domestic victimization | 1.18 (1.10, 1.27) | <0.001 |
| Polyvictimisation | 1.36 (1.09, 1.69) | 0.006 |
| **Adjusted (confounders)** |  |  |
| Total violence exposure | 1.09 (1.05, 1.14) | <0.001 |
| Witnessed community violence | 1.08 (1.00, 1.16) | 0.041 |
| Community victimization | 1.22 (1.06, 1.40) | 0.005 |
| Witnessed domestic violence | 1.53 (1.25, 1.89) | <0.001 |
| Domestic victimization | 1.15 (1.07, 1.24) | <0.001 |
| Polyvictimisation | 1.27 (1.01, 1.58) | 0.038 |
| **Adjusted (confounders and CBCL total score at age 3.5 years)** |  |  |
| Total violence exposure | 1.09 (1.05, 1.13) | <0.001 |
| Witnessed community violence | 1.07 (0.99, 1.15) | 0.080 |
| Community victimization | 1.21 (1.05, 1.39) | 0.007 |
| Witnessed domestic violence | 1.52 (1.23, 1.87) | <0.001 |
| Domestic victimization | 1.15 (1.06, 1.24) | <0.001 |
| Polyvictimisation | 1.25 (1.00, 1.56) | 0.050 |

*Note.* Based on imputed data ( $n=974$ ). Confounders include child sex, child ancestry, monthly family income, maternal education, maternal work status, maternal marital status, maternal prenatal alcohol use and smoking, child HIV exposure status, maternal antenatal depression, and child age at the 8-year CECV assessment. *Key:* CBCL=Child Behaviour Checklist; CI=confidence interval; OR=odds ratio.

### Longitudinal associations between violence exposure at 4.5 years and child psychopathology at 8 years

Table 4 presents the unadjusted and adjusted associations between violence exposure scores at 4.5 years and mental health problems at 8 years. In fully adjusted analyses, total violence exposure scores at 4.5 years were positively associated with 8-year total problem scores (B for each one-unit increase in violence score=0.27 [95% CI 0.03, 0.52], *p*=0.030), as well as with internalizing (0.24 [0.04. 0.44], *p*=0.021), and externalizing scores (0.23 [0.008, 0.45], *p*=0.042). The domestic victimization subscale score was also positively associated with all three problem scores at age 8, but there was no evidence of effects for the other types of violence exposure or polyvictimisation (see Table 4).

**Table 4.** Longitudinal associations between violence exposure score at age 4.5 years and mental health problems at age 8 years.

|  | Total CBCL Score |  |  | Internalizing CBCL Score |  |  | Externalizing CBCL Score |  |  |
| --- | --- | --- | --- | --- | --- | --- | --- | --- | --- |
|  | B (95% CI) | SE | *p* | B (95% CI) | SE | *p* | B (95% CI) | SE | *p* |
| **Unadjusted** |  |  |  |  |  |  |  |  |  |
| Total violence exposure | 0.42 (0.21, 0.64) | 0.11 | <0.001 | 0.44 (0.26, 0.62) | 0.09 | <0.001 | 0.37 (0.17, 0.56) | 0.10 | <0.001 |
| Witnessed community violence | 0.43 (0.11, 0.76) | 0.17 | 0.009 | 0.47 (0.21, 0.73) | 0.13 | <0.001 | 0.40 (0.09, 0.71) | 0.16 | 0.013 |
| Community victimization | -0.26 (-2.13, 1.60) | 0.95 | 0.782 | 0.07 (-1.32, 1.45) | 0.70 | 0.926 | -0.46 (-2.02, 1.10) | 0.79 | 0.565 |
| Witnessed domestic violence | 0.60 (-0.37, 1.56) | 0.49 | 0.228 | 1.03 (0.28, 1.77) | 0.38 | 0.007 | 0.48 (-0.32, 1.28) | 0.41 | 0.241 |
| Domestic victimization | 1.30 (0.69, 1.90) | 0.31 | <0.001 | 1.06 (0.56, 1.57) | 0.26 | <0.001 | 1.09 (0.57, 1.62) | 0.27 | <0.001 |
| Polyvictimisation | 1.30 (0.43, 2.18) | 0.45 | 0.004 | 1.40 (0.68, 2.13) | 0.37 | <0.001 | 1.13 (0.37, 1.90) | 0.39 | 0.004 |
| **Adjusted (confounders)** |  |  |  |  |  |  |  |  |  |
| Total violence exposure | 0.30 (0.06, 0.54) | 0.12 | 0.013 | 0.26 (0.06, 0.45) | 0.10 | 0.009 | 0.25 (0.04, 0.47) | 0.11 | 0.022 |
| Witnessed community violence | 0.26 (-0.10, 0.62) | 0.18 | 0.151 | 0.20 (-0.08, 0.48) | 0.14 | 0.168 | 0.25 (-0.09, 0.59) | 0.17 | 0.156 |
| Community victimization | -0.61 (-2.53, 1.31) | 0.98 | 0.535 | -0.24 (-1.62, 1.14) | 0.70 | 0.731 | -0.80 (-2.41, 0.81) | 0.82 | 0.331 |
| Witnessed domestic violence | 0.08 (-0.92, 1.08) | 0.51 | 0.871 | 0.48 (-0.27, 1.24) | 0.38 | 0.211 | 0.006 (-0.82, 0.83) | 0.42 | 0.988 |
| Domestic victimization | 1.10 (0.49, 1.71) | 0.31 | <0.001 | 0.79 (0.31, 1.28) | 0.25 | 0.001 | 0.92 (0.37, 1.46) | 0.28 | 0.001 |
| Polyvictimisation | 0.78 (-0.21, 1.78) | 0.51 | 0.123 | 0.70 (-0.10, 1.50) | 0.41 | 0.085 | 0.63 (-0.20, 1.46) | 0.42 | 0.136 |
| **Adjusted (confounders and CBCL total score at age 3.5 years)** |  |  |  |  |  |  |  |  |  |
| Total violence exposure | 0.27 (0.03, 0.52) | 0.12 | 0.030 | 0.24 (0.04, 0.44) | 0.10 | 0.021 | 0.23 (0.008, 0.45) | 0.11 | 0.042 |
| Witnessed community violence | 0.22 (-0.15, 0.59) | 0.19 | 0.240 | 0.17 (-0.12, 0.46) | 0.15 | 0.255 | 0.21 (-0.13, 0.56) | 0.18 | 0.226 |
| Community victimization | -0.65 (-2.56, 1.26) | 0.97 | 0.504 | -0.27 (-1.63, 1.08) | 0.69 | 0.693 | -0.83 (-2.46, 0.79) | 0.83 | 0.313 |
| Witnessed domestic violence | 0.08 (-0.92, 1.08) | 0.51 | 0.870 | 0.48 (-0.28, 1.24) | 0.39 | 0.212 | 0.006 (-0.82, 0.83) | 0.42 | 0.988 |
| Domestic victimization | 1.03 (0.42, 1.65) | 0.31 | 0.001 | 0.75 (0.25, 1.24) | 0.25 | 0.003 | 0.86 (0.32, 1.41) | 0.28 | 0.002 |
| Polyvictimisation | 0.66 (-0.34, 1.66) | 0.51 | 0.194 | 0.61 (-0.19, 1.41) | 0.41 | 0.136 | 0.53 (-0.30, 1.37) | 0.42 | 0.210 |

*Note.* Based on imputed data ( $n=974$ ). Robust (Huber-White) SEs were used to address some heteroscedasticity. Confounders include child sex, child ancestry, monthly family income, maternal education, maternal work status, maternal marital status, maternal prenatal alcohol use and smoking, child HIV exposure status, and maternal antenatal depression. *Key:* CBCL=Child Behaviour Checklist; CI=confidence interval.

Table 5 presents the unadjusted and adjusted associations between childhood violence exposure scores at age 4.5 years and the presence of any psychiatric disorder at age 8 years. Fully adjusted analyses found no evidence that the presence of any psychiatric disorder at age 8 was predicted by total violence exposure scores at 4.5 years (adjusted OR=1.00 [95% CI 0.93, 1.06], *p*=0.922), or by any of the violence subscales or polyvictimisation.

**Table 5.** Longitudinal associations between violence exposure score at age 4.5 years and psychiatric disorders at age 8 years.

|  | Any disorder |  |
| --- | --- | --- |
|  | OR (95% CI) | *p* |
| **Unadjusted** |  |  |
| Total violence exposure | 1.01 (0.95, 1.07) | 0.754 |
| Witnessed community violence | 0.98 (0.89, 1.07) | 0.586 |
| Community victimization | 1.27 (0.88, 1.82) | 0.203 |
| Witnessed domestic violence | 1.11 (0.90, 1.38) | 0.328 |
| Domestic victimization | 1.06 (0.93, 1.21) | 0.391 |
| Polyvictimisation | 1.07 (0.85, 1.35) | 0.549 |
| **Adjusted (confounders)** |  |  |
| Total violence exposure | 1.00 (0.94, 1.07) | 0.931 |
| Witnessed community violence | 0.97 (0.88, 1.07) | 0.493 |
| Community victimization | 1.15 (0.80, 1.66) | 0.441 |
| Witnessed domestic violence | 1.10 (0.87, 1.38) | 0.432 |
| Domestic victimization | 1.05 (0.91, 1.21) | 0.486 |
| Polyvictimisation | 1.06 (0.82, 1.37) | 0.662 |
| **Adjusted (confounders and CBCL total score at age 3.5 years)** |  |  |
| Total violence exposure | 1.00 (0.93, 1.06) | 0.922 |
| Witnessed community violence | 0.96 (0.87, 1.06) | 0.406 |
| Community victimization | 1.15 (0.80, 1.65) | 0.461 |
| Witnessed domestic violence | 1.10 (0.87, 1.39) | 0.410 |
| Domestic victimization | 1.04 (0.90, 1.20) | 0.614 |
| Polyvictimisation | 1.04 (0.80, 1.35) | 0.778 |

*Note.* Based on imputed data ( $n=974$ ). Confounders include child sex, child ancestry, monthly family income, maternal education, maternal work status, maternal marital status, maternal prenatal alcohol use and smoking, child HIV exposure status, and maternal antenatal depression. *Key:* CBCL=Child Behaviour Checklist; CI=confidence interval; OR=odds ratio.

### Sensitivity analyses

In sensitivity analyses, there was some evidence of sex differences in the cross-sectional models only. Specifically, violence exposure-by-sex interaction effects were observed for: total violence and externalizing problems; witnessed domestic violence and total, internalizing, and externalizing problems; and polyvictimisation and externalizing problems. In all cases, sex-stratified analyses evidenced greater impact of violence exposure on psychopathology in boys compared to girls (see Appendix 9).

## DISCUSSION

In this unique South African birth cohort, we report cross-sectional and longitudinal associations between violence exposure and mental health in school-aged children, including diagnosed psychiatric disorders. Strikingly, more than 90% of children had been exposed to violence by age 8 years, and 53.6% had experienced more than one type of violence. Several key findings emerged. First, given that most children had experienced domestic or community violence, the prevalence of mental health problems was relatively low in this sample, with lower-than-average CBCL scores and 11.4% meeting criteria for any psychiatric disorder. Second, while some longitudinal associations were observed between violence exposure by 4.5 years and mental health problems at 8 years, these were present only for overall violence exposure and domestic victimization scores, and only when CBCL scores were the outcome rather than diagnosable disorders. Third, in contrast, we observed strong cross-sectional associations between almost all types of violence exposure and CBCL problem scores, as well as increased odds of meeting diagnostic criteria for at least one psychiatric disorder, at age 8. Finally, there was some evidence of sex differences, such that exposure to certain types of violence had a greater impact on psychopathology in boys compared to girls.

South Africa has one of the highest rates of violence in the world,^34^ as shown by our study findings in which most children had been exposed to some form of violence by age 8. We found robust evidence that violence exposure is associated with poorer mental health in a representative sample of school-aged children. In cross-sectional analyses, total violence exposure and subtypes of witnessing community violence, witnessing domestic violence, and domestic victimization were each positively associated with caregiver-reported psychopathology on the CBCL, with similar associations for total problem, internalizing, and externalizing scores. We also showed, to our knowledge for the first time, equivalent cross-sectional associations between violence exposure and mental disorder established via structured clinical interview with primary school-aged children, with every one-point increase in overall violence exposure being associated with a 9% increase in the odds of psychiatric disorder. This observation is important, given that previous cohort studies have tended to rely solely on caregiver reports of both violence exposure and children’s mental health, leading to concerns that single-informant bias could be inflating associations.^35^ The low prevalence of mental disorders in this cohort precluded the investigation of links between violence exposure and specific disorder classes, which, alongside the use of structured diagnostic interviews with children, should be a key focus for future research.

Despite robust cross-sectional associations, we observed weaker evidence of longitudinal associations between violence exposure in the pre-school period and mental health at age 8. Only total violence exposure and domestic victimization scores at 4.5 years were positively related to 8-year CBCL scores, and we found no evidence that earlier violence exposure was predictive of psychiatric disorders at age 8. These findings are striking given hypotheses in the literature about the particular importance of adversity occurring in the first 5-years of life, a period of exceptionally rapid neural, cognitive, and social development during which there is presumed to be significant potential for biological embedding of stress responding.^36,37^ We found much more widespread evidence of longitudinal associations between violence and mental health in unadjusted analyses, highlighting the importance of birth cohort studies in addressing potential sociodemographic confounding in this field. Nonetheless, our findings suggest that in this cohort, while violence exposure in the first five years demonstrated some long-term effects on child mental health, more recent experiences of violence among primary school-aged children are substantially more important, which is consistent with parallel observations obtained in a large cohort of Brazilian adolescents.^35^ It is possible that ongoing participation in these cohorts may itself act as a protective factor for children and adolescents through active research and referral pathways for mental health and other difficulties. This may be especially true in LMICs given that public healthcare systems are often under-resourced and over-subscribed.^13^

In terms of violence subtypes, domestic victimization emerged as the most consistent predictor of mental health problems and was a common experience in this context, affecting 53.9% of children by age 8. Childhood domestic victimization is a particularly key concern in South Africa, where a significant proportion of child homicides are related to child maltreatment (44.5%).^38^ Nonetheless, each one-unit increase in the domestic victimization score at age 8 was associated with a 15% increase in the odds of any psychiatric disorder at age 8. Our findings consolidate previous evidence of the impact of domestic victimization on the mental health of children living in LMICs,^17,39–41^ and reinforce the need for early prevention and intervention strategies that effectively reduce exposure to domestic victimization among pre-school and school-aged children. A recent review highlighted the success of both parenting and community-based interventions in reducing violence against children,^42^ although implementation of these interventions is far from straightforward.

Violence against children is more common in countries with social norms that tolerate violence,^43^ with previous evidence showing that some forms of domestic victimization, including harsh parenting and corporal punishment, remain relatively acceptable in South Africa.^44^ Additionally, violence against children is typically more common in countries with high levels of inequality,^43^ with South Africa previously ranked as the most unequal of 164 countries examined globally.^45^ Such risk factors may result in challenges in establishing prevention and intervention strategies, and even when this is possible, efforts can be limited by these same circumstances.^46^ Parenting and community-based interventions may therefore be insufficient in LMICs, with changes needing to occur in an integrated way across societal, political, and legal levels to maximise sustained effects.

We also found some further, tentative evidence for differential effects of specific violence types on mental health at age 8. Witnessing domestic violence showed the strongest associations with poor mental health outcomes, with every one-unit increase being associated with a 51% increase in the odds of any psychiatric disorder at age 8 years. By contrast, though witnessing community violence was the most common exposure in our sample, with 74.7% of children exposed by 8 years, it tended to show weaker associations with mental health problems. Rates of witnessing community violence were primarily driven by two events, frequently rated as being experienced few or many times: heard gunshots (56.9% exposed at age 8) and seeing someone being beaten up (51.2%). The frequent occurrence of these events for South African children could indicate some level of desensitisation,^5,47,48^ such that they were perceived as less threatening and their subsequent impact on mental health is weaker, or they may be so ubiquitous that individual level effects become difficult to discern. Finally, the weakest evidence we observed was in relation to community victimization, which was only cross-sectionally related to increased odds of any psychiatric disorder at age 8. Despite this overall pattern in the current sample indicating that violence experienced in the home is likely to be more detrimental to children’s mental health compared to community violence, it is also notable that all forms of violence exposure contributed to poorer wellbeing in a dose-dependent fashion, consistent with existing reviews on the impact of polyvictimisation.^8,9^

A final key finding was of potential sex differences in the consequences of violence exposure. Consistent with previous evidence,^49^ including in South Africa,^23^ boys experienced greater violence exposure compared to girls in our sample (see Appendix 7). We found that certain types of violence exposure had a greater negative impact on the mental health of boys compared to girls. Total violence, witnessing domestic violence, and polyvictimisation scores were all more strongly associated with externalizing problems in boys compared to girls.

Surprisingly, witnessing domestic violence was also more strongly associated with internalizing problems in boys compared to girls. Further investigation of possible sex differences is warranted given that stereotyped gender roles and norms contribute to children’s gender socialization and play a critical role in children’s beliefs, attitudes, and behaviours, with significant implications for violence prevention efforts.^50^

This study addressed several limitations of the existing evidence base. We extended previous work to investigate violence-mental health associations among school-aged children in South Africa. Our study builds upon a small number of existing longitudinal studies focused on children living in LMICs, which have tended to focus on specific victimization types (e.g., school victimization), and/or specific high-risk groups (e.g., orphans, high-conflict areas, and HIV-affected). We used prospective data to examine both cross-sectional and longitudinal associations across multiple types of violence exposure in a representative birth cohort sample. This is also the first LMIC study, to our knowledge, to use diagnostic data from structured clinical interviews with children, addressing the potential for single informant bias.

We adjusted for a range of baseline confounders, including maternal substance use and indicators of socioeconomic status, as well as for child mental health problems at age 3.5 years which enabled us to examine whether violence exposure predicted mental health problems over and above any psychopathology present earlier in childhood. Furthermore, our violence exposure scores reflect both the number and frequency of exposures. Previous research has typically used a cumulative approach, where the number of events is tallied, and therefore equates events such as prolonged physical abuse to a one-off accident, for instance. Our approach ensures that repeated exposure to violent events is captured and accounted for.

However, several limitations must also be considered. Firstly, most of our confounders were assessed in the antenatal period, and some will have changed during the study. Secondly, there was an absence of detailed temporal information, and it is possible that the onset of mental health problems may have preceded, and possibly increased the risk of, exposure to violence, particularly for cross-sectional analyses.^41^ Third, there were some changes in caregivers that occurred over time, which could have affected our ability to detect longitudinal associations, although we note that all caregivers had significant child contact and robust effects should not rely on a single informant. Fourth, caregiver reports may underestimate child violence exposure, with social desirability biases being problematic for reports of violence occurring within the home, and caregivers potentially having limited knowledge of their children’s community experiences. Collecting violence exposure reports from children themselves in a sensitive and developmentally appropriate way should be a priority for future research. Finally, the low prevalence of psychiatric disorders in our sample limited our ability to investigate associations for internalizing and externalizing disorders separately; further investigation is therefore warranted in future waves of this cohort.

In sum, although we found that violence exposure was highly prevalent among school-aged children in South Africa, mental health symptoms and disorders were relatively uncommon at age 8 years. In combination, our strong cross-sectional associations but weaker longitudinal evidence suggest that more recent experiences of violence may be more important for children’s mental health, and again highlight the methodological limitations of retrospective, cross-sectional evidence from adults. Potential sex differences warrant further investigation given the influence of stereotypical gender roles and norms in the socialization of children, and the general societal acceptance of violence against both women and children in many LMICs, including South Africa. Future research should prioritize collecting self-reports from children and adolescents, including the use of structured diagnostic interviews, and on further investigating the longitudinal impact of exposure to different types of violence on youth in LMICs.

## CRediT STATEMENT

**Conceptualization:** MB, GH, GF, DJS, KD, SLH. **Formal analysis:** MB, GH, RS.

**Investigation:** LT, NH, HJZ, DJS, KD. **Resources:** NH, TB. **Writing – Original**

**Draft:** MB, SLH. **Writing – Review and Editing:** MB, GH, GF, LT, NH, TB, RS, AD, CA, HJZ, KD, SLH. **Supervision:** GH, GF, KD, SLH. **Project administration:** NH, TB, RS, HJZ, DJS, KD. **Funding acquisition:** MB, GF, NH, AD, CA, HJZ, DJS, KD, SLH.

## DATA AVAILABILITY

The DCHS is committed to the principle of data sharing. De-identified data will be made available to requesting researchers as appropriate. Requests for collaborations are welcome. More information can be found on the DCHS website: http://www.paediatrics.uct.ac.za/scah/dclhs. The analysis code used in this study is available through a public GitHub repository at https://github.com/megan-l-bailey/DCHS-Violence-MH.

## ACKNOWLEDGEMENTS

The authors thank the mothers and children of the Drakenstein Child Health Study who continue to participate in the cohort, as well as the clinical, administrative, and study staff of the Western Cape Government Health Department at Paarl Hospital and at the Mbekweni and TC Newman clinics for their continued support of the cohort.

