## Appendices for "Violence Exposure and Mental Health Problems Among School-Aged Children in a South African Birth Cohort"

### Supplementary Appendices

**Appendix 1.** Linearity of polyvictimisation

Polyvictimisation was captured by coding the number of subscales in which a child had experienced a traumatic event, with a maximum score of 4 indicating that the child had been exposed to events across all four subscales. We used likelihood-ratio tests to confirm linearity in polyvictimisation-psychopathology associations by comparing models capturing polyvictimisation as a categorical exposure and models capturing polyvictimisation as a numeric (continuous) exposure. Results are presented in Appendix 1b. One likelihood-ratio test was significant (the unadjusted longitudinal model for ‘any psychiatric disorder’; *p*=0.026). Given all other tests were non-significant (as well as the fully adjusted version of this model), we proceeded with capturing polyvictimisation as a numeric (continuous) exposure in all our models.

**Appendix 1b.** Likelihood-ratio tests examining the linearity of the relationship between polyvictimisation and psychopathology at age 8

| | $\chi^2$ | *p* |
|---|---|---|
| **Unadjusted: Longitudinal (polyvictimisation at 4.5 years)** | | |
| Total CBCL score | 2.15 | 0.542 |
| Internalizing CBCL score | 3.40 | 0.335 |
| Externalizing CBCL score | 2.84 | 0.417 |
| Any psychiatric disorder (MINI) | 9.31 | 0.026 |
| **Adjusted: Longitudinal (polyvictimisation at 4.5 years)** | | |
| Total CBCL score | 2.19 | 0.535 |
| Internalizing CBCL score | 1.91 | 0.591 |
| Externalizing CBCL score | 3.78 | 0.286 |
| Any psychiatric disorder (MINI) | 5.15 | 0.162 |
| **Unadjusted: Cross-sectional (polyvictimisation at 8 years)** | | |
| Total CBCL score | 1.36 | 0.714 |
| Internalizing CBCL score | 2.12 | 0.548 |
| Externalizing CBCL score | 2.58 | 0.460 |
| Any psychiatric disorder (MINI) | 5.83 | 0.120 |
| **Adjusted: Cross-sectional (polyvictimisation at 8 years)** | | |
| Total CBCL score | 1.17 | 0.759 |
| Internalizing CBCL score | 1.45 | 0.695 |
| Externalizing CBCL score | 1.96 | 0.580 |
| Any psychiatric disorder (MINI) | 4.87 | 0.181 |

*Note*. The degrees of freedom was 3 for all analyses. *Key:* CBCL=Child Behavior Checklist; MINI=Mini International Neuropsychiatric Interview.

**Appendix 2.** Open Science Framework (OSF) pre-registration

This study was pre-registered on the Open Science Framework on May 14th, 2025. We were unable to carry out some of our analysis plans. Firstly, alongside examining the association between violence exposure and 'any psychiatric disorder' (assessed using the Mini International Neuropsychiatric Interview for Children and Adolescents),[1] we planned to examine this association separately for internalizing disorders (meeting criteria for at least one of the following disorders: generalizing anxiety disorder, obsessive compulsive disorder, posttraumatic stress disorder, separation anxiety disorder, social anxiety disorder, specific phobia, and major depressive disorder) and externalizing disorders (meeting criteria for at least one of the following disorders: attention deficit hyperactivity disorder, conduct disorder, and oppositional defiant disorder). However, few children met criteria for any of the assessed psychiatric disorders (*n*=31 for internalizing disorders and *n*=67 for externalizing disorders), and we were therefore unable to examine these associations with sufficient statistical power. Secondly, we planned to estimate the proportion of mental health problems (using the Child Behaviour Checklist; CBCL)[2] and disorders (MINI-KID) at age 8 that could be explained by any violence exposure up to age 8 using population attributable fractions. We planned to binary code violence exposure to capture exposed versus unexposed children as well as to binary code CBCL *t*-scores to capture children with clinically significant elevations. We were unable to conduct these analyses, again due to low frequencies and insufficient statistical power. The majority of children (91.1%) were violence exposed at age 8, while in contrast few children had clinically significant elevations on the CBCL (*n*=47 for the total problems score, *n*=63 for the internalizing problems score, and *n*=63 for the externalizing problems score) or met criteria for a psychiatric disorder (*n*=91). We also made a minor change to our cross-sectional models. The variation in child age at the 8-year follow-up was larger than what we were expecting, with some children as old as 10.5 years when the measures were assessed for this wave. Prior to conducting any analyses, we decided to include child age at the 8-year Child Exposure to Community Violence Checklist (CECV) assessment as a confounder in our cross-sectional models, given that we hypothesised that child age could be associated with both violence exposure scores and mental health problems. Longitudinal models were carried out as planned.

**Appendix 3.** Missing data

**Appendix 3a.** Patterns and approach

Missingness for violence exposure was 23.5% at 4.5 years and 14.2% at 8 years. Missingness for age 8 psychopathology was 16.1% for the CBCL and 15.5% for the MINI-KID. A summary of missing data for all analysis variables, confounders, and auxiliary variables used in the imputation process is presented in Appendix 3b.

A comparison of children with complete data and children with missing data for violence exposure at ages 4.5 and 8 years and/or psychopathology at age 8 are presented in Appendices 3c and 3d. Complete case analyses are valid if the analysis model outcome is unrelated to missingness, conditional on the analysis model covariates.[3] While the association between the outcome and missingness cannot be examined using observed data, we are able to examine whether this assumption is violated because of a confounder that is associated with both missingness and the analysis model outcome. Findings for analyses investigating the association between our confounders and model outcomes are presented in Appendix 3e. Child ethnicity, maternal alcohol use during pregnancy, and maternal smoking during pregnancy were all associated with at least one outcome variable and with missingness. Given that these variables were also considered confounders (decided a priori based on a directed acyclic graph), we adjusted our models for these variables to avoid bias. Child age at the 8-year CECV assessment was also significantly associated with at least one model outcome and missingness in the cross-sectional analyses and was therefore also adjusted for in our cross-sectional analyses. Additionally, the total violence exposure scores at ages 3.5 and 4.5 years were also significantly associated with our outcome variables and with missingness. Given that our violence exposure scores reflect lifetime exposure, we were unable to condition on these variables. However, violence scores across all available timepoints (3.5, 4.5, 6, and 8 years), with different patterns of missingness, were included in the imputation models as auxiliary variables to make the missing at random assumption more plausible. Finally, the CBCL total problems score at 2 years was also significantly associated with our outcome variables and with missingness in our longitudinal analyses, and this was also used as an auxiliary variable in our imputation analyses.

We used multivariate imputation by chained equations with 50 imputed datasets to address missing data (*N*=974).[4] We used Stata Version 18. Auxiliary variables included: total mental health problem scores assessed using the CBCL at 2 years, 5 years, and 6.5 years and CECV subscale scores at ages 3.5 and 6 years. Two imputation models were used. The first model imputed the CECV subscale scores at 4.5 and 8 years with all outcome, confounder, and auxiliary variables. The total CECV score was then calculated after imputation. The second model was necessary due to multicollinearity when the subscale and the polyvictimisation scores were included in the same model. We therefore passively imputed the polyvictimisation scores at 4.5 and 8 years from the subscales. Due to already capturing the subscale-outcome associations in the first imputation model, we were able to remove the individual subscales from the imputation equations for the outcomes and confounders, while including the polyvictimisation variables, which addressed multicollinearity. Convergence checks prior to imputation revealed that the community victimization score at 4.5 years perfectly predicted the 'any psychiatric disorder' variable in males; the 'augment' command was therefore added to the imputation equation for the 'any disorder' variable. All binary variables were imputed using logistic regression and all continuous variables were imputed using predictive mean matching. Variables were also imputed by sex, enabling the examination of sex differences. All Monte Carlo errors for effect estimates were less than 10% of its standard error, suggesting 50 imputed datasets was sufficient.[5] Stata do-files, which detail our imputation model inclusions and exclusions, are available open access on GitHub (https://github.com/megan-l-bailey/DCHS-Violence-MH).

**Appendix 3b.** Summary of missing data (*N*=974)

| Variable | Description | Missing values | Range | Mean | SD | Skewness | Kurtosis |
|---|---|---|---|---|---|---|---|
| **Main Analysis Variables** | | | | | | | |
| totcbcl_8y | CBCL total score at age 8 years | 157 (16.1%) | 24-80 | 45.03 | 11.82 | 0.626 | 0.011 |
| intcbcl_8y | CBCL internalizing score at age 8 years | 157 (16.1%) | 33-83 | 43.96 | 9.50 | <0.001 | 0.911 |
| extcbcl_8y | CBCL externalizing score at age 8 | 157 (16.1%) | 33-77 | 48.14 | 10.33 | <0.001 | 0.003 |
| anydis | Any MINI-KID disorder at age 8 years | 151 (15.5%) | 0/1 | - | - | - | - |
| totalcecv_54m | CECV total score at age 4.5 years | 229 (23.5%) | 0-27 | 3.55 | 3.99 | <0.001 | <0.001 |
| witcom_54m | CECV witnessed community violence at age 4.5 | 229 (23.5%) | 0-22 | 2.35 | 2.69 | <0.001 | <0.001 |
| comvic_54m | CECV community victimization at age 4.5 | 229 (23.5%) | 0-4 | 0.15 | 0.52 | <0.001 | <0.001 |
| witdom_54m | CECV witnessed domestic violence at age 4.5 | 229 (23.5%) | 0-6 | 0.46 | 0.96 | <0.001 | <0.001 |
| domvic_54m | CECV domestic victimization at age 4.5 | 229 (23.5%) | 0-11 | 0.59 | 1.46 | <0.001 | <0.001 |
| polyvic_54m | CECV polyvictimisation at age 4.5 | 229 (23.5%) | 0-4 | 1.22 | 0.99 | <0.001 | 0.817 |
| totalcecv_8y | CECV total score at age 8 years | 138 (14.2%) | 0-41 | 5.93 | 4.99 | <0.001 | <0.001 |
| witcom_8y | CECV witnessed community violence at age 8 | 138 (14.2%) | 0-16 | 2.95 | 2.91 | <0.001 | <0.001 |
| comvic_8y | CECV community victimization at age 8 | 138 (14.2%) | 0-9 | 0.58 | 1.23 | <0.001 | <0.001 |
| witdom_8y | CECV witnessed domestic violence at age 8 | 138 (14.2%) | 0-5 | 0.45 | 0.90 | <0.001 | <0.001 |
| domvic_8y | CECV domestic victimization at age 8 | 138 (14.2%) | 0-20 | 1.94 | 2.56 | <0.001 | <0.001 |
| polyvic_8y | CECV polyvictimisation at age 8 | 138 (14.2%) | 0-4 | 1.81 | 1.04 | 0.022 | <0.001 |
| **Confounders** | | | | | | | |
| sex | Child sex | 0 | 0/1 | - | - | - | - |
| ethnicity | Child ethnicity | 0 | 0/1 | - | - | - | - |
| income | Monthly family income at birth | 0 | 0/1/2 | - | - | - | - |
| education | Maternal education at birth | 0 | 0/1 | - | - | - | - |

| | | | | | | | |
|---|---|---|---|---|---|---|---|
| work | Maternal work status at birth | 0 | 0/1 | - | - | - | - |
| marital_status | Maternal marital status at birth | 0 | 0/1 | - | - | - | - |
| malcohol | Maternal prenatal alcohol consumption | 41 (4.2%) | 0/1 | - | - | - | - |
| msmoking | Maternal prenatal smoking | 0 | 0/1 | - | - | - | - |
| chiv_birth | Child HIV exposure status at birth | 0 | 0/1 | - | - | - | - |
| mdepression | Maternal antenatal depression (EPDS score) | 111 (11.4%) | 0-26 | 9.51 | 5.26 | <0.001 | 0.103 |
| age_8y | Child age at the 8-year CECV assessment | 138 (14.2%) | 7.77-10.58 | 8.46 | 0.50 | <0.001 | <0.001 |
| totcbcl_42m | CBCL total score at 3.5 years | 77 (7.9%) | 28-89 | 39.43 | 9.95 | <0.001 | 0.002 |
| **Auxiliary Variables** | | | | | | | |
| totcbcl_24m | CBCL total score at age 2 years | 342 (35.1%) | 28-85 | 45.13 | 11.88 | <0.001 | 0.385 |
| totcbcl_60m | CBCL total score at age 5 years | 123 (12.6%) | 28-86 | 41.44 | 12.34 | <0.001 | 0.135 |
| totcbcl_78m | CBCL total score at age 6.5 years | 147 (15.1%) | 24-74 | 42.38 | 10.29 | 0.010 | 0.733 |
| totalcecv_42m | CECV total score at age 4.5 years | 450 (46.2%) | 0-27 | 3.43 | 3.99 | <0.001 | <0.001 |
| witcom_42m | CECV witnessed community violence at age 3.5 | 450 (46.2%) | 0-18 | 2.15 | 2.64 | <0.001 | <0.001 |
| comvic_42m | CECV community victimization at age 3.5 | 450 (46.2%) | 0-6 | 0.14 | 0.56 | <0.001 | <0.001 |
| witdom_42m | CECV witnessed domestic violence at age 3.5 | 450 (46.2%) | 0-9 | 0.54 | 1.03 | <0.001 | <0.001 |
| domvic_42m | CECV domestic victimization at age 3.5 | 450 (46.2%) | 0-10 | 0.60 | 1.36 | <0.001 | <0.001 |
| polyvic_42m | CECV polyvictimisation at age 3.5 | 450 (46.2%) | 0-4 | 1.21 | 1.03 | <0.001 | 0.129 |
| totalcecv_6y | CECV total score at age 6 years | 439 (45.1%) | 0-46 | 3.90 | 4.68 | <0.001 | <0.001 |
| witcom_6y | CECV witnessed community violence at age 6 | 439 (45.1%) | 0-16 | 2.40 | 2.74 | <0.001 | <0.001 |
| comvic_6y | CECV community victimization at age 6 | 439 (45.1%) | 0-6 | 0.22 | 0.74 | <0.001 | <0.001 |
| witdom_6y | CECV witnessed domestic violence at age 6 | 439 (45.1%) | 0-8 | 0.40 | 1.01 | <0.001 | <0.001 |
| domvic_6y | CECV domestic victimization at age 6 | 439 (45.1%) | 0-20 | 0.87 | 1.96 | <0.001 | <0.001 |
| polyvic_6y | CECV polyvictimisation at age 6 | 439 (45.1%) | 0-4 | 1.27 | 1.06 | <0.001 | 0.472 |

*Key*: CBCL=Child Behavior Checklist; CECV=Child Exposure to Community Violence Checklist; EPDS=Edinburgh Postnatal Depression Scale; MINI-KID=Mini International Neuropsychiatric Interview for Children and Adolescents.

**Appendix 3c.** Comparison of complete cases and those with missing information for violence exposure at age 4.5 years and mental health problems at age 8 years

| **Variable** | **Proportion missingness** | **Complete Cases (*n*=545)** | | **Missing Cases (*n*=429)** | | **Association with missingness** | |
|---|---|---|---|---|---|---|---|
| | *n*(%) | *n* | (%) or M(SD) | *n* | (%) or M(SD) | OR (95% CI) | *p*-value |
| **Exposure** | | | | | | | |
| CBCL total score at 2 years | 342 (35.1%) | 376 | 46.09 (12.31) | 256 | 43.70 (11.08) | 0.98 (0.97, 1.00) | 0.013 |
| CECV total score at age 3.5 years | 450 (46.2%) | 349 | 3.80 (4.17) | 175 | 2.69 (3.51) | 0.92 (0.87, 0.97) | 0.003 |
| **Confounders** | | | | | | | |
| Female sex | 0 (0%) | 265 | (48.6%) | 214 | (49.9%) | 1.05 (0.82, 1.35) | 0.696 |
| Child ethnicity (Black African)[a] | 0 (0%) | 257 | (47.2%) | 273 | (63.6%) | 1.96 (1.51, 2.54) | <0.001 |
| Child HIV exposure (exposed, uninfected)[b] | 0 (0%) | 110 | (20.2%) | 102 | (23.8%) | 1.23 (0.91, 1.67) | 0.178 |
| Monthly family income[c] | 0 (0%) | | | | | | |
| R1000-5000/m | | 269 | (49.4%) | 242 | (56.4%) | 1.33 (1.01, 1.75) | 0.044 |
| >R5000/m | | 72 | (13.2%) | 49 | (11.4%) | 1.01 (0.66, 1.53) | 0.978 |
| Maternal education (completed secondary/any tertiary)[d] | 0 (0%) | 196 | (36.0%) | 168 | (39.2%) | 1.15 (0.88, 1.49) | 0.306 |
| Maternal employment status (working)[e] | 0 (0%) | 133 | (24.4%) | 122 | (28.4%) | 1.23 (0.92, 1.64) | 0.155 |
| Maternal marital status (married/cohabiting)[f] | 0 (0%) | 219 | (40.2%) | 170 | (39.6%) | 0.98 (0.75, 1.27) | 0.860 |
| Maternal prenatal alcohol use (yes) | 41 (4.2%) | 93 | (17.1%) | 27 | (7.0%) | 0.36 (0.23, 0.57) | <0.001 |
| Maternal prenatal smoking (yes) | 0 (0%) | 198 | (36.3%) | 89 | (20.8%) | 0.46 (0.34, 0.61) | <0.001 |
| Maternal antenatal depression | 111 (11.4%) | 545 | 9.45 (5.32) | 318 | 9.62 (5.16) | 1.01 (0.98, 1.03) | 0.660 |
| CBCL total score at 3.5 years | 77 (7.9%) | 545 | 39.84 (10.35) | 352 | 38.79 (9.28) | 0.99 (0.98, 1.00) | 0.124 |

*Note*. [a]Reference group is mixed ancestry. [b]Reference group is HIV unexposed. [c]Reference group is <R1000/m. [d]Reference group is primary/some secondary. [e]Reference group is not working. [f]Reference group is single. *Key:* CBCL=Child Behavior Checklist; CECV=Child Exposure to Community Violence Checklist; CI=confidence interval; M=mean; OR=odds ratio; SD=standard deviation.

**Appendix 3d.** Comparison of complete cases and those with missing information for violence exposure and mental health problems at age 8 years

| **Variable** | **Proportion missingness** | **Complete Cases (*n*=659)** | | **Missing Cases (*n*=315)** | | **Association with missingness** | |
|---|---|---|---|---|---|---|---|
| | *n*(%) | *n* | (%) or M(SD) | *n* | (%) or M(SD) | OR (95% CI) | *p*-value |
| **Exposure** | | | | | | | |
| CBCL total score at age 2 years | 342 (35.1%) | 447 | 45.62 (12.09) | 185 | 43.94 (11.28) | 0.99 (0.97, 1.00) | 0.106 |
| CBCL total score at age 5 years | 123 (12.6%) | 604 | 41.48 (12.27) | 247 | 41.36 (12.52) | 1.00 (0.99, 1.01) | 0.905 |
| CBCL total score at age 6.5 years | 147 (15.1%) | 565 | 42.48 (10.22) | 262 | 42.15 (10.45) | 1.00 (0.98, 1.01) | 0.662 |
| CECV total score at age 3.5 years | 450 (46.2%) | 397 | 3.67 (4.07) | 127 | 2.68 (3.66) | 0.93 (0.87, 0.99) | 0.016 |
| CECV total score at age 4.5 years | 229 (23.5%) | 545 | 3.81 (4.11) | 200 | 2.87 (3.54) | 0.93 (0.89, 0.98) | 0.005 |
| CECV total score at age 6 years | 439 (45.1%) | 417 | 4.01 (4.88) | 118 | 3.50 (3.86) | 0.97 (0.93, 1.02) | 0.297 |
| **Confounders** | | | | | | | |
| Female sex | 0 (0%) | 323 | (49.0%) | 156 | (49.5%) | 1.02 (0.78, 1.34) | 0.882 |
| Child ethnicity (Black African)[a] | 0 (0%) | 320 | (48.6%) | 210 | (66.7%) | 2.12 (1.60, 2.80) | <0.001 |
| Child HIV exposure (exposed, uninfected)[b] | 0 (0%) | 129 | (19.6%) | 83 | (26.4%) | 1.47 (1.07, 2.02) | 0.017 |
| Monthly family income[c] | 0 (0%) | | | | | | |
| R1000-5000/m | | 327 | (49.6%) | 184 | (58.4%) | 1.38 (1.03, 1.86) | 0.032 |
| >R5000/m | | 89 | (13.5%) | 32 | (10.2%) | 0.88 (0.55, 1.41) | 0.600 |
| Maternal education (completed secondary/any tertiary)[d] | 0 (0%) | 242 | (36.7%) | 122 | (38.7%) | 1.09 (0.83, 1.44) | 0.545 |
| Maternal employment status (working)[e] | 0 (0%) | 163 | (24.7%) | 92 | (29.2%) | 1.26 (0.93, 1.70) | 0.138 |
| Maternal marital status (married/cohabiting)[f] | 0 (0%) | 263 | (39.9%) | 126 | (40.0%) | 1.00 (0.76, 1.32) | 0.978 |
| Maternal prenatal alcohol use (yes) | 41 (4.2%) | 99 | (15.0%) | 21 | (7.7%) | 0.47 (0.29, 0.77) | 0.003 |
| Maternal prenatal smoking (yes) | 0 (0%) | 225 | (34.1%) | 62 | (19.7%) | 0.47 (0.34, 0.65) | <0.001 |

| | | | | | | | |
|---|---|---|---|---|---|---|---|
| Maternal antenatal depression | 111 (11.4%) | 659 | 9.42 (5.28) | 204 | 9.83 (5.18) | 1.01 (0.99, 1.05) | 0.328 |
| Child age at the 8-year CECV assessment | 138 (14.2%) | 659 | 8.42 (0.43) | 177 | 8.61 (0.69) | 2.05 (1.49, 2.81) | <0.001 |
| CBCL total score at 3.5 years | 77 (7.9%) | 659 | 39.70 (10.23) | 238 | 38.66 (9.10) | 0.99 (0.97, 1.00) | 0.164 |

*Note*. [a]Reference group is mixed ancestry. [b]Reference group is HIV unexposed. [c]Reference group is <R1000/m. [d]Reference group is primary/some secondary. [e]Reference group is not working. [f]Reference group is single. *Key:* CBCL=Child Behavior Checklist; CECV=Child Exposure to Community Violence Checklist; CI=confidence interval; M=mean; OR=odds ratio; SD=standard deviation.

**Appendix 3e.** Associations between confounders and model outcomes at age 8 years

| | **CBCL Total Score** | | **CBCL Internalizing Score** | | **CBCL Externalizing Score** | | **Any Psychiatric Disorder** | |
|---|---|---|---|---|---|---|---|---|
| | B (95% CI) | *p* | B (95% CI) | *p* | B (95% CI) | *p* | OR (95% CI) | *p* |
| **Confounders** | | | | | | | | |
| Female sex | -3.17 (-4.97, -1.37) | 0.001 | -3.72 (-5.15, -2.30) | <0.001 | -2.67 (-4.25, -1.08) | 0.001 | 0.21 (0.11, 0.39) | <0.001 |
| Child ethnicity (Black African)[a] | -2.03 (-3.84, -0.22) | 0.028 | -2.95 (-4.39, -1.51) | <0.001 | -1.87 (-3.46, -0.28) | 0.022 | 1.00 (0.61, 1.64) | 0.996 |
| Child HIV exposure (exposed, uninfected)[B] | 0.008 (-2.28, 2.30) | 0.995 | -0.75 (-2.59, 1.08) | 0.420 | -0.42 (-2.44, 1.59) | 0.682 | 1.27 (0.71, 2.29) | 0.424 |
| Monthly family income (R1000-5000/m)[c] | 0.65 (-1.33, 2.62) | 0.520 | 0.38 (-1.20, 1.97) | 0.634 | 0.65 (-1.09, 2.39) | 0.460 | 0.72 (0.42, 1.23) | 0.230 |
| Monthly family income (>R5000/m) [c] | 0.62 (-2.27, 3.51) | 0.673 | 0.73 (-1.59, 3.04) | 0.539 | 0.30 (-2.25, 2.84) | 0.819 | 1.17 (0.58, 2.39) | 0.660 |
| Maternal education (completed secondary/any tertiary)[d] | -0.15 (-2.03, 1.73) | 0.874 | -1.04 (-2.55, 0.47) | 0.176 | -0.23 (-1.89, 1.42) | 0.781 | 1.17 (0.71, 1.94) | 0.532 |
| Maternal employment status (working)[e] | 0.18 (-1.93, 2.28) | 0.869 | -0.68 (-2.36, 1.01) | 0.432 | 0.09 (-1.77, 1.94) | 0.927 | 0.99 (0.56, 1.75) | 0.973 |
| Maternal marital status (married/cohabiting)[f] | 1.72 (-0.13, 3.57) | 0.068 | 1.58 (0.10, 3.07) | 0.036 | 1.61 (-0.02, 3.24) | 0.053 | 1.15 (0.70, 1.89) | 0.583 |
| Maternal prenatal alcohol use (yes) | 3.36 (0.83, 5.88) | 0.009 | 3.60 (1.58, 5.62) | <0.001 | 2.50 (0.27, 4.73) | 0.028 | 1.23 (0.64, 2.35) | 0.538 |
| Maternal prenatal smoking (yes) | 3.39 (1.49, 5.28) | <0.001 | 2.83 (1.31, 4.35) | <0.001 | 2.96 (1.29, 4.63) | 0.001 | 1.17 (0.70, 1.94) | 0.556 |
| Maternal antenatal depression | 0.29 (0.12, 0.46) | 0.001 | 0.22 (0.09, 0.36) | 0.001 | 0.24 (0.09, 0.39) | 0.002 | 1.01 (0.97, 1.06) | 0.587 |

| | | | | | | | | |
|---|---|---|---|---|---|---|---|---|
| Child age at the 8-year CECV assessment | 0.58 (-1.54, 2.71) | 0.591 | 1.73 (0.02, 3.43) | 0.047 | 1.20 (-0.67, 3.07) | 0.208 | 0.65 (0.36, 1.20) | 0.169 |
| CBCL total score at 3.5 years | 0.16 (0.08, 0.25) | <0.001 | 0.13 (0.06, 0.20) | <0.001 | 0.13 (0.06, 0.21) | 0.001 | 1.03 (1.01, 1.05) | 0.008 |
| **Auxiliary Variables** | | | | | | | | |
| CBCL total score at age 2 years | 0.22 (0.13, 0.31) | <0.001 | 0.15 (0.08, 0.22) | <0.001 | 0.19 (0.11, 0.27) | <0.001 | 1.01 (0.99, 1.04) | 0.241 |
| CBCL total score at age 5 years | 0.20 (0.12, 0.28) | <0.001 | 0.11 (0.05, 0.18) | <0.001 | 0.18 (0.11, 0.25) | <0.001 | 1.02 (1.00, 1.04) | 0.105 |
| CBCL total score at age 6.5 years | 0.34 (0.25, 0.43) | <0.001 | 0.18 (0.11, 0.26) | <0.001 | 0.32 (0.24, 0.40) | <0.001 | 1.06 (1.03, 1.09) | <0.001 |
| CECV total score at age 3.5 years | 0.73 (0.43, 1.02) | <0.001 | 0.66 (0.43, 0.90) | <0.001 | 0.67 (0.41, 0.93) | <0.001 | 1.09 (1.02, 1.16) | 0.006 |
| CECV total score at age 4.5 years | 0.47 (0.23, 0.71) | <0.001 | 0.45 (0.26, 0.65) | <0.001 | 0.38 (0.17, 0.60) | <0.001 | 1.02 (0.96, 1.08) | 0.536 |
| CECV total score at age 6 years | 0.51 (0.27, 0.74) | <0.001 | 0.45 (0.27, 0.63) | <0.001 | 0.41 (0.20, 0.62) | <0.001 | 1.04 (0.99, 1.10) | 0.094 |

*Note*. For confounder associations, analyses are based on complete case data for the total violence exposure score at age 8, mental health problems at age 8, and confounders (*n*=659). For auxiliary variable associations, analyses are additionally based on complete data for each of the variables of interest (*n*=447 for CBCL total at 2 years; *n*=604 for CBCL total at 5 years; *n*=565 for CBCL total at 6.5 years; *n*=397 for CECV total at 3.5 years; *n*=545 for CECV total at 4.5 years; and *n*=417 for CECV total at 6 years). [a]Reference group is mixed ancestry. [b]Reference group is HIV unexposed. [c]Reference group is <R1000/m. [d]Reference group is primary/some secondary. [e]Reference group is not working. [f]Reference group is single. Key: B=unstandardized beta; CBCL=Child Behavior Checklist; CECV=Child Exposure to Community Violence Checklist; CI=confidence interval; OR=odds ratio.

**Appendix 4.** Complete case analyses

**Appendix 4a.** Sociodemographic characteristics, trauma exposure, and mental health outcome descriptives

| | Complete Cases | *n* (%) | Mean (SD) | Range |
|---|---|---|---|---|
| **Sociodemographic characteristics** | | | | |
| Child sex (female) | 974 | 479 (49.2%) | | |
| Child ethnicity (Black African)[a] | 974 | 530 (54.4%) | | |
| Child HIV exposure (exposed but uninfected)[b] | 974 | 212 (21.8%) | | |
| Monthly family income | 974 | | | |
| <R1000 | | 342 (35.1%) | | |
| R1000-5000 | | 511 (52.5%) | | |
| >R5000 | | 121 (12.4%) | | |
| Maternal education (completed secondary/any tertiary)[c] | 974 | 364 (37.4%) | | |
| Maternal employment status (working)[d] | 974 | 255 (26.2%) | | |
| Maternal marital status (married/cohabiting)[e] | 974 | 389 (39.9%) | | |
| Maternal prenatal alcohol use (yes) | 933 | 120 (12.9%) | | |
| Maternal prenatal tobacco use (yes) | 974 | 287 (29.5%) | | |
| Maternal antenatal depression | 863 | | 9.51 (5.26) | 0-26 |
| Child age at the 8-year CECV assessment | 836 | | 8.46 (0.50) | 7.77-10.58 |
| **Child trauma exposure – CECV scores at 4.5 years** | | | | |
| Total | 745 | 559 (75.0%) | 3.55 (3.99) | 0-27 |
| Witnessed community violence | 745 | 496 (66.6%) | 2.35 (2.69) | 0-22 |
| Community victimization | 745 | 67 (9.0%) | 0.15 (0.52) | 0-4 |
| Witnessed domestic violence | 745 | 178 (23.9%) | 0.46 (0.96) | 0-6 |
| Domestic victimization | 745 | 168 (22.6%) | 0.59 (1.46) | 0-11 |
| Polyvictimisation | 745 | | 1.22 (0.99) | 0-4 |

| | | | | |
|---|---|---|---|---|
| 0 | | 186 (25.0%) | | |
| 1 | | 309 (41.5%) | | |
| 2 | | 166 (22.3%) | | |
| 3 | | 68 (9.1%) | | |
| 4 | | 16 (2.2%) | | |
| **Child trauma exposure – CECV scores at 8 years** | | | | |
| Total | 836 | 760 (90.9%) | 5.93 (4.99) | 0-41 |
| Witnessed community violence | 836 | 635 (76.0%) | 2.95 (2.91) | 0-16 |
| Community victimization | 836 | 230 (27.5%) | 0.58 (1.23) | 0-9 |
| Witnessed domestic violence | 836 | 204 (24.4%) | 0.45 (0.90) | 0-5 |
| Domestic victimization | 836 | 442 (52.9%) | 1.94 (2.56) | 0-20 |
| Polyvictimisation | 836 | | 1.81 (1.04) | 0-4 |
| 0 | | 76 (9.1%) | | |
| 1 | | 277 (33.1%) | | |
| 2 | | 260 (31.1%) | | |
| 3 | | 178 (21.3%) | | |
| 4 | | 45 (5.4%) | | |
| **Child mental health** | | | | |
| CBCL total score at 3.5 years | 897 | | 39.43 (9.95) | 28-89 |
| CBCL total score at 8 years | 817 | | 45.03 (11.82) | 24-80 |
| CBCL internalizing score at 8 years | 817 | | 43.96 (9.50) | 33-83 |
| CBCL externalizing score at 8 years | 817 | | 48.14 (10.33) | 33-77 |
| Any psychiatric disorder at 8 years | 823 | 91 (11.1%) | | |

*Note*. Full sample demographics and descriptive statistics (*N*=974). [a]Reference group is mixed ancestry. [b]Reference group is HIV unexposed. [c]Reference group is primary/some secondary. [d]Reference group is not working. [e]Reference group is single. *Key*: CBCL=Child Behavior Checklist; CECV=Child Exposure to Community Violence Checklist.

**Appendix 4b.** Cross-sectional associations between trauma exposure score at age 8 years and mental health problems at age 8 years

| | Total CBCL Score | | | Internalizing CBCL Score | | | Externalizing CBCL Score | | |
|---|---|---|---|---|---|---|---|---|---|
| | B (95% CI) | SE | *p* | B (95% CI) | SE | *p* | B (95% CI) | SE | *p* |
| **Unadjusted** | | | | | | | | | |
| Overall violence exposure | 0.58 (0.39, 0.77) | 0.10 | <0.001 | 0.41 (0.23, 0.58) | 0.09 | <0.001 | 0.52 (0.35, 0.69) | 0.09 | <0.001 |
| Witnessed community violence | 0.61 (0.32, 0.89) | 0.15 | <0.001 | 0.56 (0.32, 0.80) | 0.12 | <0.001 | 0.55 (0.29, 0.82) | 0.14 | <0.001 |
| Community victimization | 1.02 (0.15, 1.90) | 0.45 | 0.022 | 0.70 (0.01, 1.40) | 0.35 | 0.048 | 0.98 (0.24, 1.73) | 0.38 | 0.009 |
| Witnessed domestic violence | 1.81 (0.73, 2.89) | 0.55 | 0.001 | 1.33 (0.43, 2.24) | 0.46 | 0.004 | 1.49 (0.52, 2.46) | 0.49 | 0.003 |
| Domestic victimization | 0.88 (0.52, 1.25) | 0.18 | <0.001 | 0.44 (0.11, 0.77) | 0.17 | 0.010 | 0.79 (0.47, 1.11) | 0.16 | <0.001 |
| Polyvictimisation | 2.47 (1.62, 3.32) | 0.43 | <0.001 | 1.57 (0.87, 2.27) | 0.36 | <0.001 | 2.15 (1.41, 2.88) | 0.37 | <0.001 |
| **Adjusted (confounders)** | | | | | | | | | |
| Overall violence exposure | 0.48 (0.29, 0.67) | 0.10 | <0.001 | 0.29 (0.12, 0.45) | 0.09 | 0.001 | 0.44 (0.27, 0.61) | 0.09 | <0.001 |
| Witnessed community violence | 0.47 (0.16, 0.78) | 0.16 | 0.003 | 0.31 (0.05, 0.57) | 0.13 | 0.019 | 0.42 (0.13, 0.70) | 0.15 | 0.004 |
| Community victimization | 0.70 (-0.13, 1.53) | 0.42 | 0.099 | 0.40 (-0.24, 1.03) | 0.32 | 0.220 | 0.73 (0.01, 1.44) | 0.36 | 0.047 |
| Witnessed domestic violence | 1.29 (0.21, 2.37) | 0.55 | 0.019 | 0.83 (-0.03, 1.69) | 0.44 | 0.059 | 1.06 (0.09, 2.02) | 0.49 | 0.032 |
| Domestic victimization | 0.92 (0.56, 1.28) | 0.18 | <0.001 | 0.51 (0.18, 0.84) | 0.17 | 0.003 | 0.85 (0.53, 1.17) | 0.16 | <0.001 |
| Polyvictimisation | 2.06 (1.19, 2.93) | 0.44 | <0.001 | 1.16 (0.46, 1.85) | 0.35 | 0.001 | 1.83 (1.09, 2.58) | 0.38 | <0.001 |
| **Adjusted (confounders and CBCL total score at age 3.5 years)** | | | | | | | | | |
| Overall violence exposure | 0.44 (0.25, 0.63) | 0.10 | <0.001 | 0.25 (0.08, 0.42) | 0.09 | 0.004 | 0.41 (0.24, 0.58) | 0.09 | <0.001 |
| Witnessed community violence | 0.39 (0.07, 0.71) | 0.16 | 0.016 | 0.25 (-0.02, 0.51) | 0.14 | 0.068 | 0.36 (0.07, 0.65) | 0.15 | 0.016 |
| Community victimization | 0.61 (-0.22, 1.44) | 0.42 | 0.151 | 0.33 (-0.31, 0.97) | 0.33 | 0.315 | 0.66 (-0.06, 1.37) | 0.36 | 0.071 |
| Witnessed domestic violence | 1.20 (0.12, 2.28) | 0.55 | 0.029 | 0.76 (-0.11, 1.62) | 0.44 | 0.087 | 0.99 (0.02, 1.95) | 0.49 | 0.045 |
| Domestic victimization | 0.87 (0.52, 1.23) | 0.18 | <0.001 | 0.47 (0.14, 0.79) | 0.17 | 0.005 | 0.81 (0.50, 1.13) | 0.16 | <0.001 |
| Polyvictimisation | 1.94 (1.07, 2.80) | 0.44 | <0.001 | 1.05 (0.36, 1.75) | 0.35 | 0.003 | 1.74 (0.99, 2.48) | 0.38 | <0.001 |

*Note*. Based on complete case data (*n*=685). Robust (Huber-White) SEs were used to address some heteroscedasticity. Confounders include child sex, child ethnicity, monthly family income, maternal education, maternal work status, maternal marital status, maternal prenatal alcohol and tobacco use, child HIV exposure status, maternal antenatal depression, and child age at the 8-year CECV assessment. *Key:* CBCL=Child Behaviour Checklist; CI=confidence interval.

**Appendix 4c.** Cross-sectional associations between trauma exposure score at age 8 years and mental health disorders at age 8 years

| | Any disorder | |
|---|---|---|
| | OR (95% CI) | *p* |
| **Unadjusted** | | |
| Overall violence exposure | 1.11 (1.06, 1.16) | <0.001 |
| Witnessed community violence | 1.08 (1.00, 1.16) | 0.058 |
| Community victimization | 1.36 (1.17, 1.57) | <0.001 |
| Witnessed domestic violence | 1.50 (1.21, 1.88) | <0.001 |
| Domestic victimization | 1.18 (1.09, 1.28) | <0.001 |
| Polyvictimisation | 1.62 (1.28, 2.07) | <0.001 |
| **Adjusted (confounders)** | | |
| Overall violence exposure | 1.08 (1.04, 1.13) | <0.001 |
| Witnessed community violence | 1.06 (0.97, 1.16) | 0.171 |
| Community victimization | 1.23 (1.05, 1.43) | 0.009 |
| Witnessed domestic violence | 1.45 (1.15, 1.84) | 0.002 |
| Domestic victimization | 1.15 (1.06, 1.25) | 0.001 |
| Polyvictimisation | 1.41 (1.09, 1.81) | 0.008 |
| **Adjusted (confounders and CBCL total score at age 3.5 years)** | | |
| Overall violence exposure | 1.08 (1.03, 1.12) | 0.002 |
| Witnessed community violence | 1.04 (0.96, 1.14) | 0.337 |
| Community victimization | 1.22 (1.04, 1.42) | 0.014 |
| Witnessed domestic violence | 1.41 (1.11, 1.79) | 0.004 |
| Domestic victimization | 1.14 (1.05, 1.24) | 0.003 |
| Polyvictimisation | 1.36 (1.06, 1.76) | 0.017 |

*Note*. Based on complete case data (*n*=668). Confounders include child sex, child ethnicity, monthly family income, maternal education, maternal work status, maternal marital status, maternal prenatal alcohol and tobacco use, child HIV exposure status, maternal antenatal depression, and child age at the 8-year CECV assessment. *Key:* CBCL=Child Behaviour Checklist; CI=confidence interval; OR=odds ratio.

**Appendix 4d.** Longitudinal associations between trauma exposure score at age 4.5 years and mental health problems at age 8 years

| | **Total CBCL Score** | | | **Internalizing CBCL Score** | | | **Externalizing CBCL Score** | | |
|---|---|---|---|---|---|---|---|---|---|
| | B (95% CI) | SE | *p* | B (95% CI) | SE | *p* | B (95% CI) | SE | *p* |
| **Unadjusted** | | | | | | | | | |
| Overall violence exposure | 0.47 (0.23, 0.70) | 0.12 | <0.001 | 0.45 (0.26, 0.64) | 0.10 | <0.001 | 0.38 (0.17, 0.59) | 0.11 | <0.001 |
| Witnessed community violence | 0.54 (0.17, 0.91) | 0.19 | 0.004 | 0.52 (0.23, 0.81) | 0.15 | <0.001 | 0.48 (0.14, 0.82) | 0.17 | 0.006 |
| Community victimization | -1.12 (-3.09, 0.84) | 1.00 | 0.260 | -0.63 (-2.09, 0.83) | 0.74 | 0.394 | -1.25 (-2.98, 0.48) | 0.88 | 0.158 |
| Witnessed domestic violence | 0.44 (-0.53, 1.41) | 0.49 | 0.373 | 0.96 (0.18, 1.73) | 0.39 | 0.016 | 0.19 (-0.65, 1.04) | 0.43 | 0.652 |
| Domestic victimization | 1.52 (0.95, 2.08) | 0.29 | <0.001 | 1.18 (0.65, 1.70) | 0.27 | <0.001 | 1.20 (0.70, 1.71) | 0.26 | <0.001 |
| Polyvictimisation | 1.44 (0.47, 2.41) | 0.50 | 0.004 | 1.45 (0.65, 2.25) | 0.41 | <0.001 | 1.17 (0.30, 2.04) | 0.44 | 0.008 |
| **Adjusted (confounders)** | | | | | | | | | |
| Overall violence exposure | 0.27 (0.02, 0.52) | 0.13 | 0.033 | 0.22 (0.02, 0.43) | 0.10 | 0.033 | 0.22 (-0.01, 0.44) | 0.11 | 0.061 |
| Witnessed community violence | 0.29 (-0.11, 0.68) | 0.20 | 0.151 | 0.19 (-0.11, 0.50) | 0.15 | 0.210 | 0.26 (-0.10, 0.62) | 0.18 | 0.155 |
| Community victimization | -1.77 (-3.77, 0.23) | 1.02 | 0.082 | -1.10 (-2.45, 0.24) | 0.69 | 0.108 | -1.74 (-3.57, 0.09) | 0.93 | 0.062 |
| Witnessed domestic violence | -0.31 (-1.30, 0.68) | 0.50 | 0.540 | 0.22 (-0.56, 1.00) | 0.40 | 0.574 | -0.42 (-1.29, 0.45) | 0.44 | 0.341 |
| Domestic victimization | 1.22 (0.68, 1.76) | 0.27 | <0.001 | 0.88 (0.39, 1.36) | 0.25 | <0.001 | 0.97 (0.47, 1.48) | 0.26 | <0.001 |
| Polyvictimisation | 0.55 (-0.53, 1.64) | 0.55 | 0.317 | 0.50 (-0.36, 1.36) | 0.44 | 0.253 | 0.46 (-0.50, 1.41) | 0.49 | 0.347 |
| **Adjusted (confounders and CBCL total score at age 3.5 years)** | | | | | | | | | |
| Overall violence exposure | 0.23 (-0.03, 0.50) | 0.13 | 0.083 | 0.19 (-0.02, 0.41) | 0.11 | 0.078 | 0.18 (-0.05, 0.42) | 0.12 | 0.124 |
| Witnessed community violence | 0.24 (-0.17, 0.64) | 0.21 | 0.259 | 0.16 (-0.16, 0.47) | 0.16 | 0.333 | 0.22 (-0.15, 0.59) | 0.19 | 0.242 |
| Community victimization | -1.87 (-3.88, 0.14) | 1.02 | 0.068 | -1.18 (-2.48, 0.13) | 0.66 | 0.077 | -1.82 (-3.70, 0.07) | 0.96 | 0.059 |
| Witnessed domestic violence | -0.34 (-1.33, 0.66) | 0.51 | 0.510 | 0.20 (-0.58, 0.99) | 0.40 | 0.611 | -0.44 (-1.32, 0.43) | 0.45 | 0.322 |
| Domestic victimization | 1.14 (0.58, 1.69) | 0.28 | <0.001 | 0.82 (0.31, 1.32) | 0.26 | 0.002 | 0.91 (0.39, 1.43) | 0.26 | 0.001 |
| Polyvictimisation | 0.37 (-0.72, 1.46) | 0.56 | 0.505 | 0.37 (-0.50, 1.24) | 0.44 | 0.405 | 0.32 (-0.64, 1.28) | 0.49 | 0.515 |

*Note*. Based on complete case data (*n*=566). Robust (Huber-White) SEs were used to address some heteroscedasticity. Confounders include child sex, child ethnicity, monthly family income, maternal education, maternal work status, maternal marital status, maternal prenatal alcohol and tobacco use, child HIV exposure status, and maternal antenatal depression. *Key:* CBCL=Child Behaviour Checklist; CI=confidence interval; SE=standard error.

**Appendix 4e.** Longitudinal associations between trauma exposure score at age 4.5 years and mental health disorders at age 8 years

| | **Any disorder** | |
|---|---|---|
| | OR (95% CI) | *p* |
| **Unadjusted** | | |
| Overall violence exposure | 1.01 (0.95, 1.08) | 0.657 |
| Witnessed community violence | 0.99 (0.90, 1.09) | 0.857 |
| Community victimization | 0.99 (0.60, 1.63) | 0.978 |
| Witnessed domestic violence | 1.04 (0.80, 1.34) | 0.784 |
| Domestic victimization | 1.11 (0.97, 1.27) | 0.125 |
| Polyvictimisation | 1.04 (0.80, 1.35) | 0.777 |
| **Adjusted (confounders)** | | |
| Overall violence exposure | 0.99 (0.92, 1.06) | 0.727 |
| Witnessed community violence | 0.95 (0.86, 1.06) | 0.379 |
| Community victimization | 0.91 (0.56, 1.48) | 0.698 |
| Witnessed domestic violence | 0.99 (0.75, 1.31) | 0.942 |
| Domestic victimization | 1.08 (0.93, 1.24) | 0.320 |
| Polyvictimisation | 0.95 (0.71, 1.27) | 0.729 |
| **Adjusted (confounders and CBCL total score at age 3.5 years)** | | |
| Overall violence exposure | 0.98 (0.91, 1.05) | 0.529 |
| Witnessed community violence | 0.94 (0.85, 1.05) | 0.280 |
| Community victimization | 0.90 (0.55, 1.45) | 0.655 |
| Witnessed domestic violence | 0.99 (0.74, 1.31) | 0.923 |
| Domestic victimization | 1.06 (0.91, 1.23) | 0.466 |
| Polyvictimisation | 0.92 (0.69, 1.23) | 0.563 |

*Note*. Based on complete case data (*n*=561). Confounders include child sex, child ethnicity, monthly family income, maternal education, maternal work status, maternal marital status, maternal prenatal alcohol and tobacco use, child HIV exposure status, and maternal antenatal depression. *Key:* CBCL=Child Behaviour Checklist; CI=confidence interval; OR=odds ratio.

**Appendix 4f.** Sex differences

We conducted sex-specific sensitivity analyses using complete case data. Eight interaction effects were significant. The longitudinal association between community victimization score at age 4.5 years and any psychiatric disorder at age 8 significantly differed by sex (interaction effect *p*-value=0.024). Sex stratified analyses revealed that the community victimization OR in girls (adjusted OR 1.91 [95% CI 0.84, 4.35], *p*=0.120) was greater than the OR for boys (0.71 [0.37, 1.35], *p*=0.299), though associations for both sexes were non-significant. All other significant interaction effects were cross-sectional models. The associations between the total violence exposure score at age 8 and both the total problems score (interaction effect *p*-value=0.042) and the externalizing problems score (interaction effect *p*-value=0.020) significantly differed by sex. Sex stratified analyses revealed that violence exposure was more strongly associated with total (0.53 [0.27, 0.78], *p*<0.001) and externalizing problem scores (0.52 [0.29, 0.74], *p*<0.001) in boys compared to girls (total: 0.22 [-0.07, 0.51], *p*=0.132; externalizing: 0.17 [-0.08, 0.41], *p*=0.190). Furthermore, the associations between witnessed domestic violence score at age 8 and all three problem scores at age 8 significantly differed by sex (total: *p*<0.001; internalizing: *p* =0.007; externalizing: *p*=0.001). Sex stratified analyses again revealed a greater impact of witnessed domestic violence on mental health problems in boys (total: 2.80 [1.34, 4.26], *p*<0.001; internalizing: 1.59 [0.32, 2.85], *p*=0.014; externalizing: 2.20 [0.81, 3.58], *p*=0.002) compared to girls (total: -1.14 [-2.66, 0.38], *p*=0.142; internalizing: -0.45 [-1.53, 0.63], *p*=0.412; externalizing: -0.77 [-2.02, 0.47], *p*=0.224). Finally, the association between the polyvictimisation score at age 8 and both the total problems score (*p*=0.005) and the externalizing problems score (*p*<0.001) significantly differed by sex. Again, sex stratified analyses revealed a greater impact of polyvictimisation on total and externalizing problem scores in boys (total: 2.92 [1.69, 4.15], *p*<0.001; externalizing: 2.91 [1.83, 3.99], *p*<0.001) compared to girls (total: 0.62 [-0.60, 1.84]), *p*=0.320; externalizing: 0.19 [-0.81, 1.19], *p*=0.710).

**Appendix 5.** Linear regression diagnostics

Examination of the distribution of regression model residuals using histograms consistently revealed a bimodal distribution with two distinct peaks. Panels A-F in Appendix 5b present the residual distribution for the fully adjusted cross-sectional associations between violence exposure scores at age 8 and the total problem score of the CBCL at age 8 as an example of this. We observed an unexpected peak of highly negative residuals, indicating that children were scoring substantially below their predicted CBCL score based on their CECV score.

Examination of the data (in the context of the cross-sectional model linking the total violence score to the total problem score) revealed that a number of children had scored zero on the CBCL (*n*=73 [8.9%]; *t*-scores of 24 for males and 25 for females), and that these children made up the majority of those with highly negative residuals (91 children had residuals that were less than -15; of these, 58 children had a score of zero on the CBCL [63.7%]). We posited that this pattern could reflect a) resilience (despite exposure to violence, these children have very few emotional and behaviour problems), or b) measurement error. For the latter, our primary concern was regarding the informant who completed the CBCL. To be eligible to complete the CBCL for a child, a caregiver must have been living with or caring for the child for three or more days per week. This was to ensure that the informant knew the child well enough to answer the questionnaires, though it is possible that this may not be the case even with this eligibility criteria. Unfortunately, the CBCL informant was not recorded, and we were therefore unable to investigate whether the informant was related to having a CBCL score of zero. We did examine other potential predictors of this, including maternal work status at age 8, which we posited could be a good proxy for CBCL informant (if the mother was working, it was more likely that she would not be able to complete the CBCL for her child either at the clinic or via telephone). Maternal work status at age 8 was not significantly associated with having a CBCL score of zero (OR 1.52 [0.93, 2.49], *p*=0.096) or with having a highly negative residual (1.32 [0.85, 2.06], *p*=0.216). Additionally, none of the sociodemographic variables included as confounders in our models were associated with having a CBCL score of zero at age 8 years.

Unfortunately, we were unable to definitively conclude whether these zero scores on the CBCL were due to resilience or measurement error without introducing any bias into our analyses. Given that we were unable to identify any plausible proxies for the CBCL informant, and that we had explored the data and attempted to explain this pattern of findings (as presented here), we proceeded with our analyses as planned. It is worth noting that should this higher-than-expected number of zero scores on the CBCL reflect measurement error, the estimates presented in our models (specifically those using the CBCL outcomes) may underestimate the association between violence exposure and child emotional and behavioral problems. Our violence-disorder models (using the MINI-KID outcome) may provide a more accurate and unbiased picture of this association given that diagnoses were obtained via clinical interviews delivered to children (in the presence of a caregiver) by research assistants trained to conduct the interview.

**Appendix 5b.** Residual distributions for the fully adjusted cross-sectional associations between violence exposure scores and the total problem score on the CBCL at age 8 years

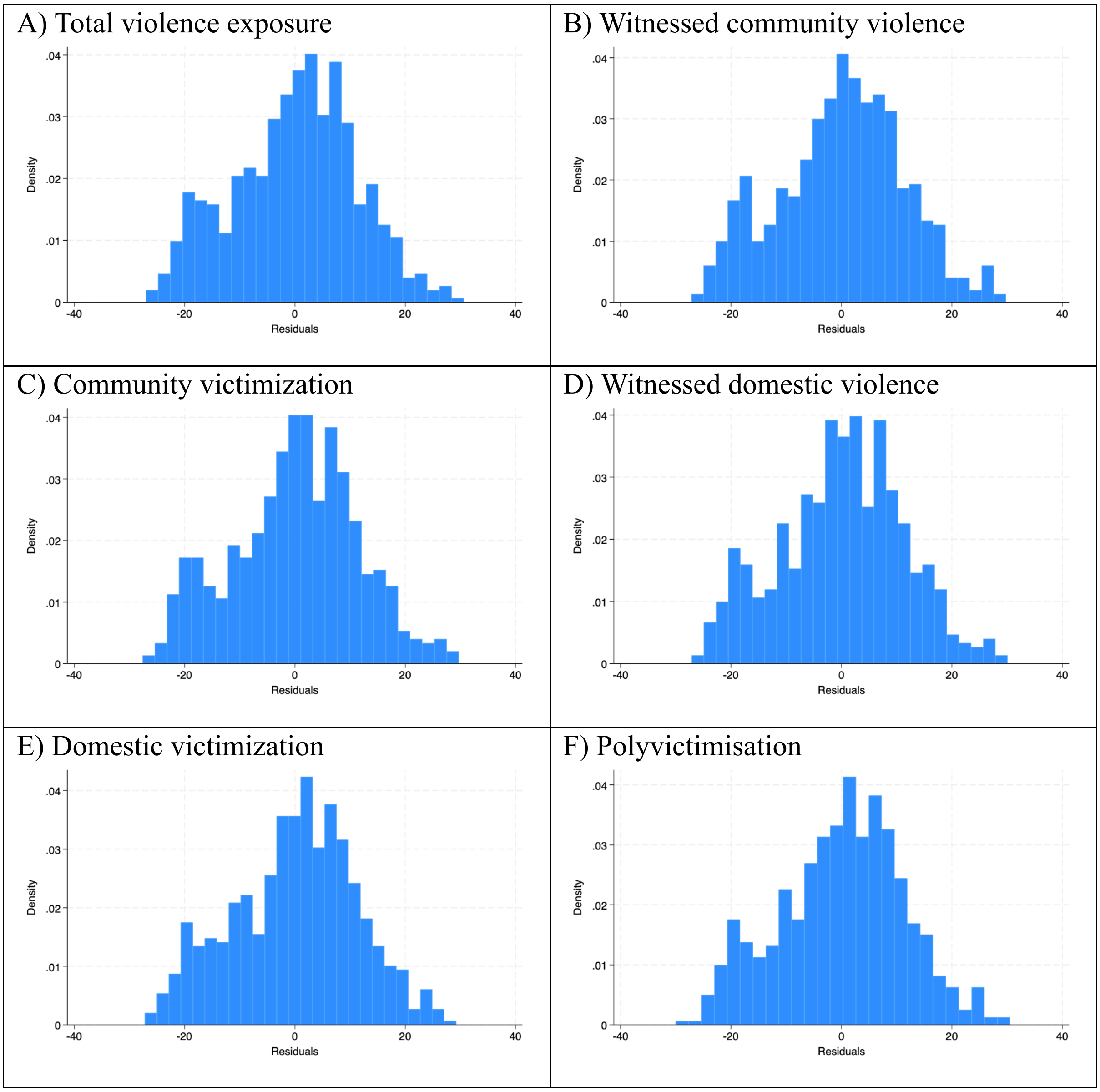

**Appendix 6.** Comparison of sociodemographic characteristics for children included versus excluded in the analysis sample

| | Total Sample | | Included (*n*=974) | | Excluded (*n*=169) | | OR or Mean Difference (95% CI) | *p*-value |
|---|---|---|---|---|---|---|---|---|
| | N | *n*(%) or M(SD) | N | *n*(%) or M(SD) | N | *n*(%) or M(SD) | | |
| Female sex | 1143 | 557 (48.7%) | 974 | 479 (49.2%) | 169 | 78 (46.2%) | 0.89 (0.64, 1.23) | 0.468 |
| Child ethnicity (Black African)[a] | 1142 | 632 (55.3%) | 974 | 530 (54.4%) | 168 | 102 (60.7%) | 1.29 (0.93, 1.81) | 0.130 |
| Child HIV exposure (exposed, uninfected)[b] | 1143 | 248 (21.7%) | 974 | 212 (21.8%) | 169 | 36 (21.3%) | 0.97 (0.65, 1.45) | 0.893 |
| Monthly family income (R1000-5000/m)[c] | 1142 | 596 (52.2%) | 974 | 511 (52.5%) | 168 | 85 (50.6%) | 1.29 (0.88, 1.91) | 0.195 |
| Monthly family income (>R5000/m)[c] | | 160 (14.0%) | | 121 (12.4%) | | 39 (23.2%) | 2.51 (1.55, 4.04) | <0.001 |
| Maternal education (completed secondary/any tertiary)[d] | 1141 | 448 (39.3%) | 974 | 364 (37.4%) | 167 | 84 (50.3%) | 1.70 (1.22, 2.36) | 0.002 |
| Maternal employment status (working)[e] | 1143 | 308 (27.0%) | 974 | 255 (26.2%) | 169 | 53 (31.4%) | 1.29 (0.90, 1.84) | 0.162 |
| Maternal marital status (married/cohabiting)[f] | 1143 | 462 (40.4%) | 974 | 389 (39.9%) | 169 | 73 (43.2%) | 1.14 (0.82, 1.59) | 0.426 |
| Maternal prenatal alcohol use (yes) | 1067 | 137 (12.8%) | 933 | 120 (12.9%) | 134 | 17 (12.7%) | 0.98 (0.57, 1.70) | 0.955 |
| Maternal prenatal smoking (yes) | 1142 | 323 (28.3%) | 974 | 287 (29.5%) | 168 | 36 (21.4%) | 0.65 (0.44, 0.97) | 0.034 |
| Maternal antenatal depression | 994 | 9.45 (5.28) | 863 | 9.51 (5.26) | 131 | 9.03 (5.38) | 0.48 (-0.49, 1.45) | 0.330 |

*Note*. [a]Reference group is mixed ancestry. [b]Reference group is HIV unexposed. [c]Reference group is <R1000/m. [d]Reference group is primary/some secondary. [e]Reference group is not working. [f]Reference group is single. *Key:* CI=confidence interval; OR=odds ratio.

**Appendix 7.** Prevalence of each traumatic event at the 4.5- and 8-year follow-ups

| | 4.5 years | | | 8 years | | |
|---|---|---|---|---|---|---|
| | **Total (n=745)** | **Boys (n=374)** | **Girls (n=371)** | **Total (n=836)** | **Boys (n=423)** | **Girls (n=413)** |
| **Any violence exposure** | **559 (75.0%)** | **286 (76.5%)** | **273 (73.6%)** | **760 (90.9%)** | **396 (93.6%)** | **364 (88.1%)** |
| **Witnessed community violence** | **496 (66.6%)** | **259 (69.3%)** | **237 (63.9%)** | **635 (76.0%)** | **345 (81.6%)** | **290 (70.2%)** |
| Heard gunshots | 313 (42.0%) | 179 (47.9%) | 134 (36.1%) | 476 (56.9%) | 266 (62.9%) | 210 (50.9%) |
| Seen someone beaten up in the neighborhood | 340 (45.6%) | 168 (44.9%) | 172 (46.4%) | 428 (51.2%) | 230 (54.4%) | 198 (47.9%) |
| Seen dead body in the neighborhood | 63 (8.5%) | 39 (10.4%) | 24 (6.5%) | 87 (10.4%) | 55 (13.0%) | 32 (7.8%) |
| Seen somebody point a gun at another in the neighborhood | 41 (5.5%) | 28 (7.5%) | 13 (3.5%) | 36 (4.3%) | 27 (6.4%) | 9 (2.2%) |
| Seen somebody get shot in the neighborhood | 21 (2.8%) | 15 (4.0%) | 6 (1.6%) | 21 (2.5%) | 9 (2.1%) | 12 (2.9%) |
| Seen somebody point a knife at another in the neighborhood | 117 (15.7%) | 67 (17.9%) | 50 (13.5%) | 121 (14.5%) | 79 (18.7%) | 42 (10.2%) |
| Seen somebody get stabbed in the neighborhood | 63 (8.5%) | 37 (9.9%) | 26 (7.0%) | 70 (8.4%) | 48 (11.4%) | 22 (5.3%) |
| Seen someone forced to do something sexual neighborhood | 0 (0%) | 0 (0%) | 0 (0%) | 1 (0.1%) | 1 (0.2%) | 0 (0%) |
| Child known someone killed by another | 29 (3.9%) | 19 (5.1%) | 10 (2.7%) | 62 (7.4%) | 39 (9.2%) | 23 (5.6%) |
| Seen someone being killed by another person elsewhere | 10 (1.3%) | 7 (1.9%) | 3 (0.8%) | 13 (1.6%) | 7 (1.7%) | 6 (1.5%) |
| **Community victimization** | **67 (9.0%)** | **39 (10.4%)** | **28 (7.6%)** | **230 (27.5%)** | **145 (34.3%)** | **85 (20.6%)** |
| House robbery child present | 26 (3.5%) | 14 (3.7%) | 12 (3.2%) | 45 (5.4%) | 26 (6.2%) | 19 (4.6%) |
| Someone threatened to beat up the child at school or creche | 13 (1.7%) | 8 (2.1%) | 5 (1.4%) | 102 (12.2%) | 70 (16.6%) | 32 (7.8%) |

| | | | | | | |
|---|---|---|---|---|---|---|
| Someone threatened to beat up the child elsewhere | 14 (1.9%) | 9 (2.4%) | 5 (1.4%) | 55 (6.6%) | 34 (8.0%) | 21 (5.1%) |
| Child been beaten up elsewhere | 21 (2.8%) | 15 (4.0%) | 6 (1.6%) | 97 (11.6%) | 63 (14.9%) | 34 (8.2%) |
| Someone elsewhere threatened to kill the child | 2 (0.3%) | 0 (0%) | 2 (0.5%) | 4 (0.5%) | 3 (0.7%) | 1 (0.2%) |
| Someone at school or creche threatened to shoot or stab the child | 0 (0%) | 0 (0%) | 0 (0%) | 4 (0.5%) | 4 (1.0%) | 0 (0%) |
| Someone elsewhere threatened to shoot or stab the child | 0 (0%) | 0 (0%) | 0 (0%) | 2 (0.2%) | 2 (0.5%) | 0 (0%) |
| Someone shot or stabbed the child elsewhere | 0 (0%) | 0 (0%) | 0 (0%) | 0 (0%) | 0 (0%) | 0 (0%) |
| **Witnessed domestic violence** | **178 (23.9%)** | **96 (25.7%)** | **82 (22.1%)** | **204 (24.4%)** | **117 (27.7%)** | **87 (21.1%)** |
| Seen grownups at home hit each other | 168 (22.6%) | 92 (24.6%) | 76 (20.5%) | 180 (21.5%) | 101 (23.9%) | 79 (19.1%) |
| Seen somebody point gun at another at home | 4 (0.5%) | 2 (0.5%) | 2 (0.5%) | 9 (1.1%) | 5 (1.2%) | 4 (1.0%) |
| Seen someone at home get stabbed | 21 (2.8%) | 11 (2.9%) | 10 (2.7%) | 27 (3.2%) | 20 (4.7%) | 7 (1.7%) |
| Seen someone at home get shot | 1 (0.1%) | 0 (0%) | 1 (0.3%) | 2 (0.2%) | 1 (0.2%) | 1 (0.2%) |
| Seen someone forced to do something sexual | 1 (0.1%) | 1 (0.3%) | 0 (0%) | 0 (0%) | 0 (0%) | 0 (0%) |
| Seen someone being killed by another person at home | 6 (0.8%) | 2 (0.5%) | 4 (1.1%) | 4 (0.5%) | 3 (0.7%) | 1 (0.2%) |
| **Domestic victimization** | **168 (22.6%)** | **104 (27.8%)** | **64 (17.3%)** | **442 (52.9%)** | **234 (55.3%)** | **208 (50.4%)** |
| Someone threatened to beat up the child at home | 14 (1.9%) | 9 (2.4%) | 5 (1.4%) | 217 (26.0%) | 111 (26.2%) | 106 (25.7%) |
| Child been beaten up at home | 15 (2.0%) | 9 (2.4%) | 6 (1.6%) | 181 (21.7%) | 88 (20.8%) | 93 (22.5%) |
| Someone at home threatened to kill the child | 2 (0.3%) | 1 (0.3%) | 1 (0.3%) | 4 (0.5%) | 3 (0.7%) | 1 (0.2%) |
| Family member threatened to shoot or stab the child | 0 (0%) | 0 (0%) | 0 (0%) | 2 (0.2%) | 2 (0.5%) | 0 (0%) |
| Someone shot or stabbed the child at home | 2 (0.3%) | 2 (0.5%) | 0 (0%) | 0 (0%) | 0 (0%) | 0 (0%) |

| | | | | | | |
|---|---|---|---|---|---|---|
| Someone made the child do something sexual | 5 (0.7%) | 3 (0.8%) | 2 (0.5%) | 10 (1.2%) | 4 (1.0%) | 6 (1.5%) |
| Family member shouts at the child fiercely and loudly | 59 (7.9%) | 38 (10.2%) | 21 (5.7%) | 67 (8.0%) | 35 (8.3%) | 32 (7.8%) |
| Anyone at home used a stick or belt or hard item to hit the child | 49 (6.6%) | 32 (8.6%) | 17 (4.6%) | 117 (14.0%) | 84 (19.9%) | 33 (8.0%) |
| Anyone at home hit the child so hard they were hurt | 25 (3.4%) | 13 (3.5%) | 12 (3.2%) | 22 (2.6%) | 14 (3.3%) | 8 (1.9%) |
| Anyone at home said the child would be sent away or kicked out | 16 (2.2%) | 8 (2.1%) | 8 (2.2%) | 75 (9.0%) | 49 (11.6%) | 26 (6.3%) |
| Anyone at home called the child horrible names | 65 (8.7%) | 40 (10.7%) | 25 (6.7%) | 116 (13.9%) | 61 (14.4%) | 55 (13.3%) |

*Note*. Based on complete case data.

**Appendix 8.** Psychiatric disorders

Child mental health disorders were assessed at age 8 years via clinical interview using the MINI-KID [1]. Ten diagnoses were assessed: generalized anxiety disorder, obsessive compulsive disorder, posttraumatic stress disorder, separation anxiety disorder, social anxiety disorder, specific phobia, major depressive disorder, attention deficit hyperactivity disorder, conduct disorder, and oppositional defiant disorder. Few children met diagnostic criteria for each of the disorders (see Appendix 8b). We therefore were only able to examine the association between violence exposure and 'any psychiatric disorder' (binary variable capturing whether a child met criteria for at least one disorder).

**Appendix 8b.** Prevalence of mental health disorders at age 8 years

| **Diagnosis** | **Prevalence, *n*(%)** |
|---|---|
| **Any psychiatric disorder** | **91/823 (11.1%)** |
| **Any internalizing disorder** | **31/836 (3.7%)** |
| Generalized anxiety disorder | 2/838 (0.2%) |
| Obsessive compulsive disorder | 8/853 (0.9%) |
| Posttraumatic stress disorder | 2/846 (0.2%) |
| Separation anxiety disorder | 2/840 (0.2%) |
| Social anxiety disorder | 2/841 (0.2%) |
| Specific phobia | 17/850 (2.0%) |
| Major depressive disorder | 2/844 (0.2%) |
| **Any externalizing disorder** | **67/838 (8.0%)** |
| Attention deficit hyperactivity disorder | 58/848 (6.8%) |
| Conduct disorder | 25/843 (3.0%) |
| Oppositional defiant disorder | 19/835 (2.3%) |

*Note*. Based on complete case data.

**Appendix 9.** Sex differences

We conducted sex-specific sensitivity analyses using imputed data. Five interaction effects, all cross-sectional models, were statistically significant. The association between the total violence exposure score at age 8 and externalizing problems at age 8 significantly differed by sex (interaction effect *p*-value=0.016). Sex stratified analyses revealed a greater impact of violence exposure on externalizing problems in boys (adjusted B=0.56 [95% CI 0.36, 0.76], *p*<0.001) compared to girls (0.25 [0.03, 0.47], *p*=0.024). Furthermore, the associations between the witnessed domestic violence score at age 8 and the total problems (interaction effect *p*-value<0.001), internalizing problems (*p*=0.002), and externalizing problems scores at age 8 (*p*=0.001) all significantly differed by sex. Sex stratified analyses revealed that witnessed domestic violence was more strongly associated with total (3.01 [1.69, 4.32], *p*<0.001), internalizing (1.88 [0.68, 3.08], *p*=0.002), and externalizing problems (2.46 [1.24, 3.69], *p*<0.001) in boys but not in girls (total: -0.65 [-2.06, 0.77], *p*=0.368; internalizing: -0.32 [-1.33, 0.69], *p*=0.529; externalizing: -0.39 [-1.61, 0.84], *p*=0.535). The association between polyvictimisation at age 8 and externalizing problems at age 8 also significantly differed by sex (interaction effect *p*-value=0.020). Sex stratified analyses revealed that greater polyvictimisation was more strongly associated with externalizing scores in boys (2.11 [1.05, 3.17], *p*<0.001), but not in girls (0.58 [-0.31, 1.48], *p*=0.202).

We also found weak evidence for five further interactions: the association between the total violence score at age 8 and total problems at age 8 (interaction effect *p*-value=0.056), witnessed community violence at age 8 and both total problems (*p*=0.050) and externalizing problems at age 8 (*p*=0.078), community victimization at age 4.5 years and any psychiatric disorder at age 8 (*p*=0.083), and witnessed domestic violence at age 4.5 years and any psychiatric disorder at age 8 (*p*=0.081). In sex stratified analyses, there was a greater impact of overall violence exposure on total problem scores in boys (B=0.56 [95% CI 0.34, 0.78], *p*<0.001) compared to girls at age 8 (0.33 [0.07, 0.59], *p*=0.013). Also, there was a greater impact of witnessed community violence on total and externalizing problems in boys (total: 0.55 [0.12, 0.99], *p*=0.013; externalizing: 0.52 [0.13, 0.92], *p*=0.009) compared to girls at age 8 (total: 0.34 [-0.09, 0.77], *p*=0.119; externalizing: 0.29 [-0.06, 0.65], *p*=0.108). Surprisingly, greater community victimization by age 4.5 years was more strongly associated with meeting criteria for any psychiatric disorder at age 8 in girls (1.92 [0.93, 3.95], *p*=0.077) compared to boys (1.01 [0.66, 1.55], *p*=0.955), though both were non-significant. Similarly, greater witnessed domestic violence by 4.5 years was also more strongly associated with any psychiatric disorder at age 8 in girls (1.40 [0.96, 2.04], *p*=0.078) compared to boys (0.98 [0.73, 1.31], *p*=0.870), though both were again non-significant.
